# EIN-RECAP: Evidence-Grounded Multi-Agent Summarization and Retrieval of Infectious-Disease Expert Discussions

**DOI:** 10.64898/2026.11.05.26367381

**Authors:** Akash Choudhuri, Yongjian Zhong, Susan Beekmann, Jacob Hodges, Kishlay Jha, Alberto M. Segre, Bijaya Adhikari, Philip M. Polgreen

## Abstract

Infectious disease experts exchange practical knowledge through clinical discussions, but identifying relevant information across years of conversations is difficult. These discussions often contain differing opinions and emerging observations that are not yet reflected in published evidence. We present EIN-RECAP, an AI-assisted platform developed with Emerging Infections Network (EIN) experts and deployed in beta to summarize and search historical discussions. The system combines retrieval-grounded summarization, citation validation, and hybrid search while distinguishing published evidence from community experience. We evaluated summarization methods across 3,185 threads and conducted blinded expert review on a recent subset. Although a multi-agent workflow achieved stronger automated scores, experts favored a simpler generation pipeline and identified repetition, inaccuracies, and overconfident recommendations in the more complex system. This feedback motivated an ensemble that improved discussion coverage and key clinical points on nine high-disagreement threads, although retrieval relevance remained a limitation. Our findings demonstrate the importance of expert involvement in clinical AI development and show why automated evaluation alone is insufficient to guide system selection.

## Introduction

In 1992, the Institute of Medicine published a report (Oaks Jr, Shope, and Lederberg 1992) that identified a lack of communication between clinical medicine and public health as a threat to identifying and controlling emerging and reemerging pathogens. At the time, surveillance was only performed for specific diseases. One of the recommendations suggested the creation of multiple sentinel networks for disease surveillance. Consequently, in 1996, the Centers for Disease Control and Prevention (CDC) and the Infectious Diseases Society of America (IDSA) launched the Emerging Infections Network (EIN) to help detect and respond to infectious disease threats by connecting frontline professionals who work to treat, prevent, and control the spread of emerging and re-emerging infectious diseases to each other and to members of the public health community. Over time, the EIN has evolved into a flexible, nationwide network of ID clinicians with members from all 50 states, the District of Columbia, and Puerto Rico. Members primarily include ID physicians, but the network has expanded to include PharmDs and other Advanced Practice Providers (APPs) who work in the field of infectious diseases. In addition to front-line healthcare professionals, the network’s listserv also includes CDC investigators and other members of the public health community. As of 2014, it included more than 3,000 members of the infectious-diseases community (Pillai et al. 2014).

Through its restricted-access listserv, EIN members exchange clinical questions and experiences that may not yet be represented in guidelines or peer-reviewed publications. The data used in this work contains 6,085 threads spanning February 11, 1997, to March 4, 2026, each consisting of an initial post or announcement and its associated replies. Naively querying relevant discussions from the sheer volume of data evolving over three decades faces two primary challenges.

First, finding and bringing together discussions from the distant past requires accounting for differences in terminology and clinical context. Across an archive spanning nearly three decades, the same condition may appear under different names or abbreviations. Keyword matching can miss clinically related threads that use different terminology while returning discussions that share terms but address different questions. A recurring clinical question may also be spread across discussions from several periods. Even after locating a relevant thread, one may need to read a detailed case description and exchange to understand its conclusions. In such scenarios, summaries would help members assess relevance before committing to a full review.

Second, EIN administrators prepare monthly digests that require reviewing discussions and their replies. The work involves clinical interpretation as well as reading, since recommendations may reflect different institution-specific practices. Precomputed thread summaries could provide a consistent starting point for digest preparation. They could also support administrators in tracking recurring concerns and identifying questions that warrant formal surveys. Their value for digest preparation remains to be evaluated.

These needs call for both retrieval and summarization. Appropriately adapted large language models (LLMs) have shown strong performance on several clinical summarization tasks (Van Veen et al. 2024), while query reformulation and semantic representations can complement lexical retrieval when related questions use different terminology. Summarizing expert discussions nevertheless requires care: a generated account may omit minority viewpoints, combine recommendations made under incompatible assumptions, or present anecdotal experience as established evidence. Retrieval-augmented generation (RAG) can ground outputs in identifiable sources, but retrieved material still requires relevance screening and citation validation (Lewis et al. 2020; Zakka et al. 2024). A useful summary must distinguish published evidence from community observations and preserve the uncertainty needed to interpret recommendations.

We present **EIN-RECAP**, an **E**merging **I**nfections **N**etwork **R**etrieval and **E**vidence-grounded **C**linical **A**nalysis **P**latform for summarizing and searching historical EIN discussions. Its Multi-Agent Summarizer produces structured summaries that record clinical claims and recommendations together with discussion limitations. Claims are labeled according to support from retrieved publications, prior EIN discussions, or neither, and cited identifiers are validated against retrieved sources. Within this framework, Similar-Thread Retrieval supplies related earlier discussions as context, with temporal restrictions intended to prevent later discussions from being used to support an earlier thread.

The Multi-Agent Search Tool retrieves summaries and original discussions from natural-language clinical queries using lexical and semantic matching with clinical terminology normalization. This supports access to differently worded discussions, while the summaries help users decide which complete threads to review. Summarization and evidence validation occur offline, allowing the resulting structured outputs to be reused for online search and potential administrative workflows. Developed in collaboration with EIN experts, EIN-RECAP is deployed in beta within the network.

Our primary contributions are as follows:

- We introduce **EIN-RECAP**, an applied AI platform for searching historical EIN discussions through structured summaries linked to original threads and supporting evidence.
- We integrate evidence-grounded summarization and retrieval of related earlier discussions with terminology-aware search, separating offline summary generation and evidence validation from online retrieval. The precomputed summaries support relevance assessment and provide reusable material for archive review.
- We evaluate summarization pipelines on 3,185 threads with at least three replies and conduct blinded expert assessment on 30 recent threads, where the pipeline with the strongest ROUGE and BERTScore results received the lowest expert ratings. Expert feedback informed an ensemble redesign that improved discussion coverage and key clinical point scores on nine high-disagreement threads, while publication relevance remained a limitation across pipelines.

## Proposed System Design

Our overall framework consists of two primary components, namely the **Multi-Agent Summarizer** and **Multi-Agent Search Tool**. While the search tool enables access to the processed discussion archive, the primary methodological contribution of our framework is the Multi-Agent Summarizer. We will elaborate on the two components in detail next.

### Multi-Agent Summarizer

EIN threads often contain information that typically comes from varied perspectives from members with different levels of expertise. A single language model’s response may omit less prominent viewpoints or may sometimes highlight clinician experience as widely accepted clinical evidence even when it may simply be subjective opinions about a clinical topic. To address these limitations, our proposed frameworks assign distinct responsibilities to specialized agents and aggregate all the important information in producing a grounded summary. We will describe the two versions of this part of the overall framework, with a focus on Version 1 (V1) in the main text, while Version 2 (V2) will be described in detail in the Appendix.

#### Version 1 (V1)

V1 is a retrieval-grounded summarization pipeline with explicit evidence tracking and deterministic post-generation validation. We adopt the general retrieval-augmented generation (RAG) framework (Lewis et al. 2020) with a single generation agent, chosen deliberately for this deployment, since fewer model calls make an error attributable to a specific stage, retrieval or generation. Additionally, as prior work (Xiong et al. 2024) has demonstrated the potential value of RAG in medical applications, we curate the retrieval corpus instead of searching the unrestricted literature.

V1 consists of five connected modules, namely PubMed Retrieval, Similar Thread Retrieval, Grounded Generation, and Output and Provenance Recording. The complete processing path and the outputs at each stage are shown in Figure 1. PubMed retrieval establishes the available scientific evidence, similar-thread retrieval contributes historical community experience, grounded generation synthesizes the two without collapsing their roles, deterministic validation checks the resulting attributions, and provenance recording preserves the complete processing history.

**Figure 1:**
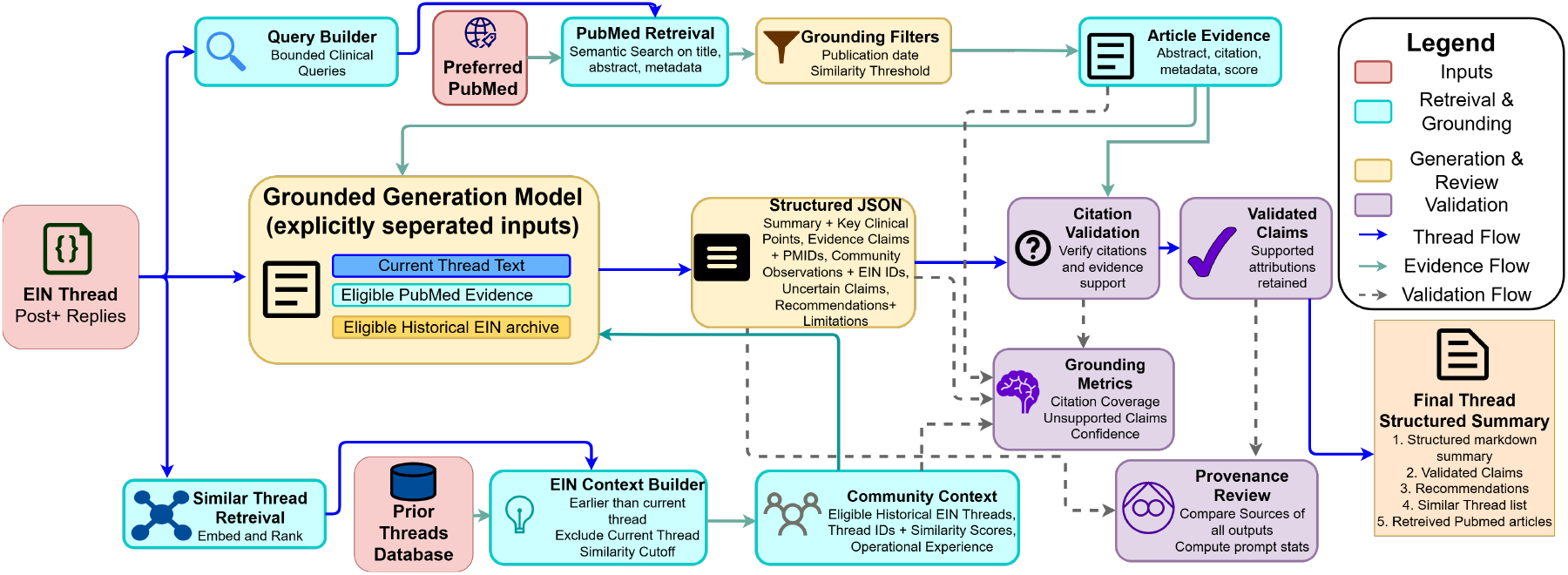
The V1 summarization pipeline. PubMed retrieval and similar-thread retrieval supply date-filtered evidence to a single grounded generation agent, which receives the thread text, published evidence, and prior EIN discussions as separate inputs and returns structured JSON. Cited identifiers are then validated deterministically against the retrieved sources, with unsupported claims reassigned to the uncertain category

**Figure 2:**
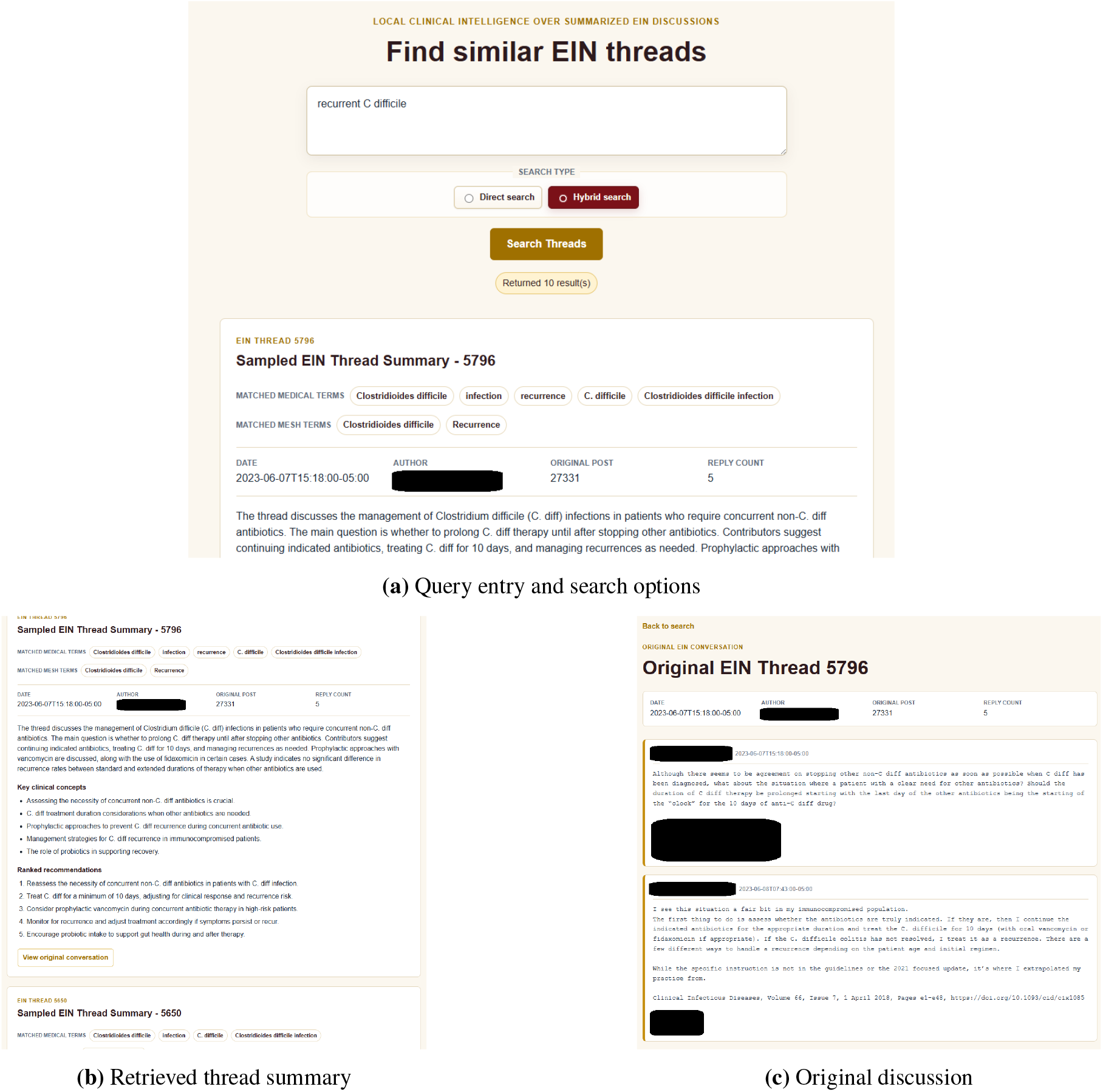
EIN-RECAP beta interface. (a) Users enter a clinical query and select direct or hybrid search. (b) Results display matched terminology, a thread summary, key clinical concepts, and recommendations. (c) Users can open the original discussion to review its context. Screenshots show selected interface regions. We have redacted any personal identifiable information.

**Figure 3:**
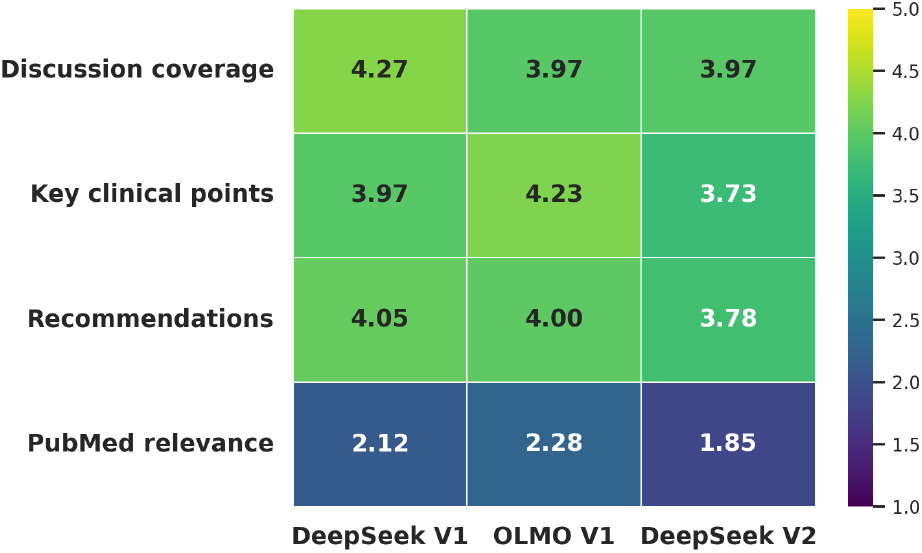
Mean Scores by Two Expert Annotators

**Figure 4:**
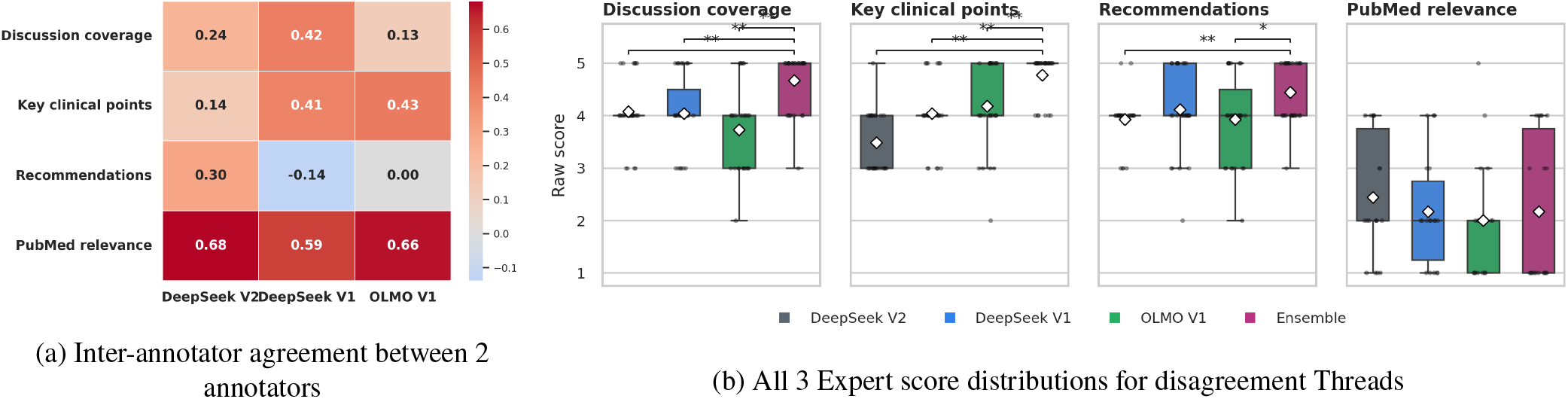
Human expert evaluation: (a) Inter-annotator agreement between 2 annotators and (b) Raw score distributions from three human expert annotators for nine high-disagreement threads, comparing the ensemble model with the three standalone LLM systems

**Figure 5:**
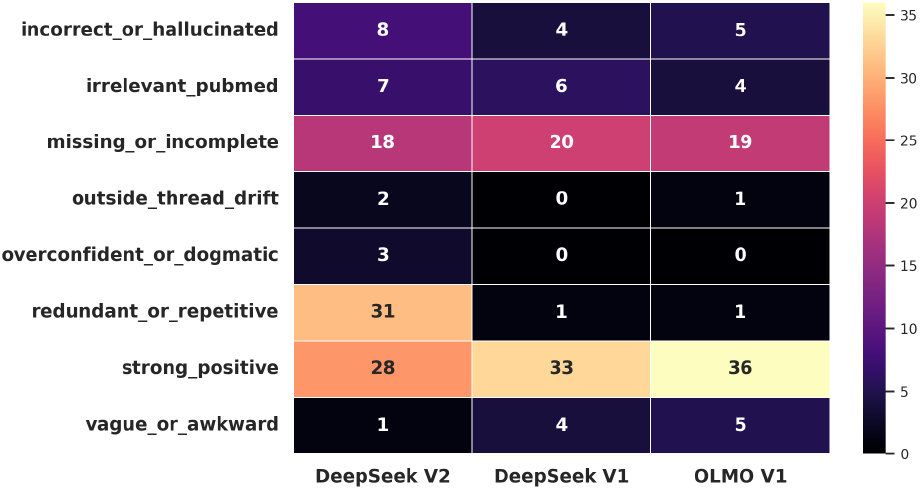
Counts of annotator free-text comments per normalized theme and standalone model

#### Preferred PubMed Retrieval Module

This module provides the information from published works related to the thread being summarized. Instead of retrieving from unrestricted biomedical literature (Fiorini et al. 2018), we first construct a local PubMed collection from a set of journals selected in consultation with our EIN collaborators. The detailed list of the journals is presented in Table 6 in the Appendix. This decision was made to obtain information from credible and relevant sources. However, this approach comes with a tradeoff, wherein potentially relevant research articles from venues outside the preferred list will not be included.

For each thread, the query builder (LLM agent) generates a set of retrieval queries. Additionally, we limit the supplied context (i.e., number of PubMed articles), as LLMs are known to use information inconsistently as the input context becomes longer (Liu et al. 2024). The resulting queries are then used to search a vector database constructed from the preferred PubMed collection. For real-time summarization, for threads originating in year *Y*_*i*_, eligible PubMed articles are restricted to publications from years *Y*_*i*_ *−*11 through *Y*_*i*_ *−*1. This, in turn, means that the works published after the discussion cannot be used to explain or retrospectively strengthen its claims. The resulting set is then passed to the generation model.

#### Similar-Thread Retrieval Module

Although PubMed provides access to peer-reviewed evidence, the published works may not address time-sensitive clinical questions that arise during an emerging outbreak. Clinical observations and studies typically require some time before being indexed into PubMed. In this situation, the experience of EIN members is extremely crucial as they have real-time knowledge of emerging conditions and topics through observation (Pillai et al. 2014), and prior discussions between EIN members in threads can provide a valuable source of clinical information that may not yet be represented in the published literature. To account for this scenario, this module retrieves semantically similar historical EIN threads and provides them to the summarization model as a separate source of community-practice evidence. Similar to PubMed retrieval, the temporal restriction prevents information from later discussions from being introduced into summaries of earlier threads.

#### Grounded Generation Module

This module synthesizes three explicitly separated inputs, namely the resolved text of the current thread, the eligible PubMed articles, and the eligible historical EIN discussions. The module’s output contains a concise thread summary, key clinical points, evidence-supported claims, community-supported observations, uncertain claims, recommendations, limitations, cited PubMed PMIDs, and cited EIN thread identifiers.

Evidence-supported claims must cite one or more PubMed articles supplied by the retrieval module. Similarly, community-supported observations must cite eligible historical EIN threads. Statements that cannot be adequately supported by either source are assigned to the uncertain-claims category. The framework also computes summary-level grounding and retrieval-confidence metrics.

These serve two goals: **1:** They make grounding failures quantifiable during evaluation. **2:** They provide diagnostic signals for identifying whether a weak summary originated from insufficient retrieval or from generation behavior.

#### Output and Provenance Recording

The final module converts the validated outputs from the individual modules into two outputs. First, the module creates a human-readable markdown summary. Second, the system records metadata that was used to generate the final human-readable artifacts. This allows us to audit the specific modules of the overall framework in case of inaccuracies.

#### Version 2 (V2)

Instead of generating a single summary directly from the thread, V2 first produces multiple candidate interpretations of the same discussion, evaluates them, and synthesizes the final structured output.

It has a set of perspective-specific Generation Agents. Each agent receives the same discussion thread, but is assigned a different clinical objective. The agent outputs are then passed to a Reflection Agent, which evaluates whether each candidate is faithful to the source conversation and other aspects. In addition, the Proximity Agent identifies related prior EIN discussions and detects similarity among generated recommendations, and the Ranking Agent compares recommendations according to their relevance. These comparisons are aggregated to produce an ordered recommendation list. Finally, an Evolution Agent refines the highest-ranked recommendations, and a Meta-review Agent integrates the outputs of the preceding agents into the final structured summary. The overall framework is presented in Figure 6 in the Appendix.

**Figure 6:**
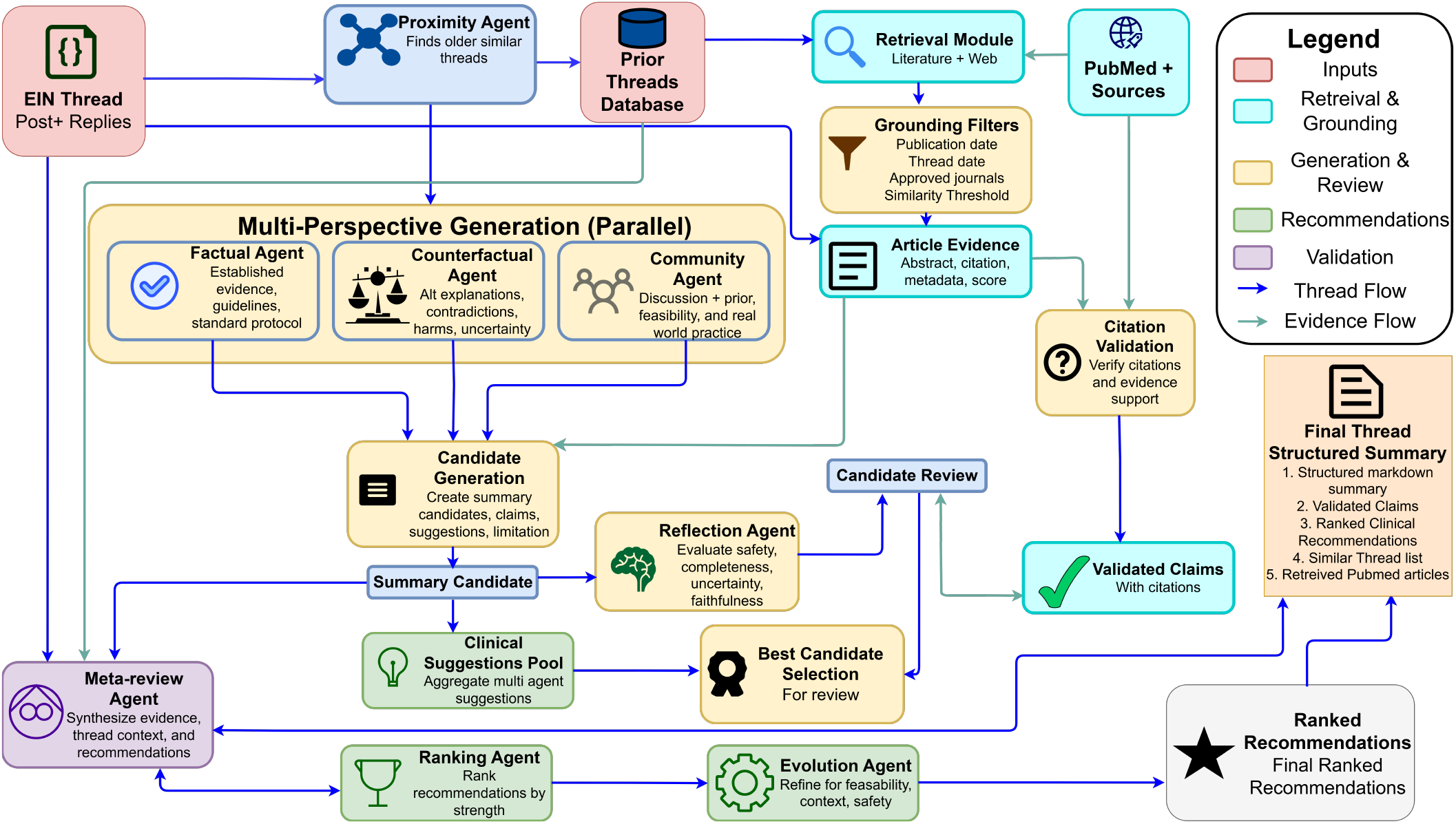
The V2 summarization pipeline. Three generation agents process the same thread in parallel under different objectives, namely factual, counterfactual, and community practice, so that clinically relevant minority interpretations survive into the candidate set. The Reflection Agent reviews each candidate for faithfulness, completeness, and treatment of uncertainty, while the Proximity Agent supplies related prior threads and detects near-duplicate recommendations. Recommendations pooled across all three generation agents are ordered by the Ranking Agent using pairwise Elo comparison and refined by the Evolution Agent. The Meta-review Agent then combines the candidate with the strongest reflection score, which supplies the summary prose and clinical concepts, with the ranked recommendation list, so a recommendation can reach the output even when the candidate it came from was not selected. Citation validation remains deterministic, as in V1.

**Figure 7:**
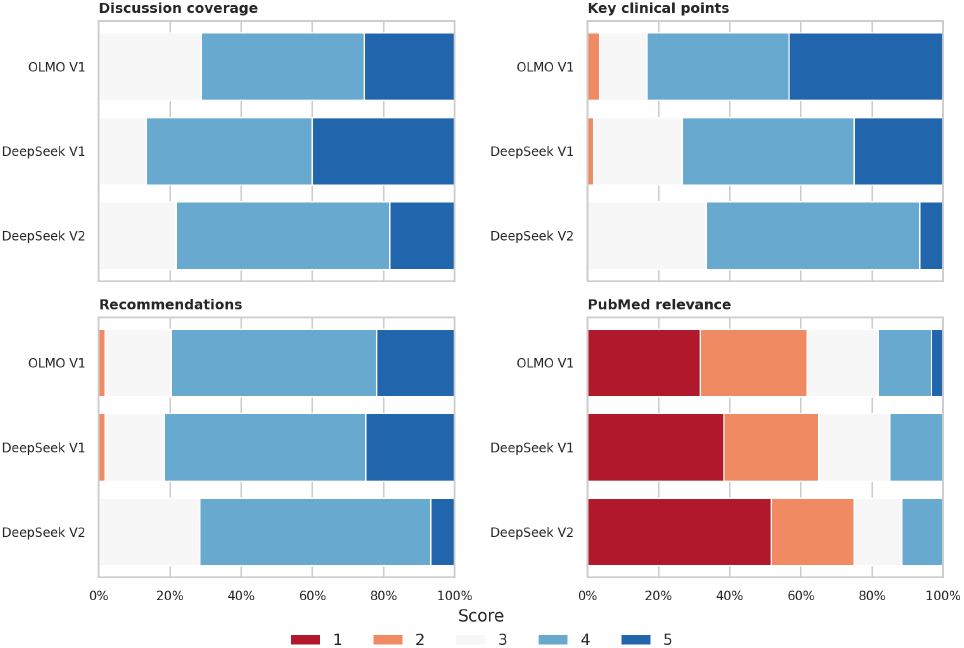
Raw Score Distribution Among the Two Expert Annotators for the Threads.

**Figure 8:**
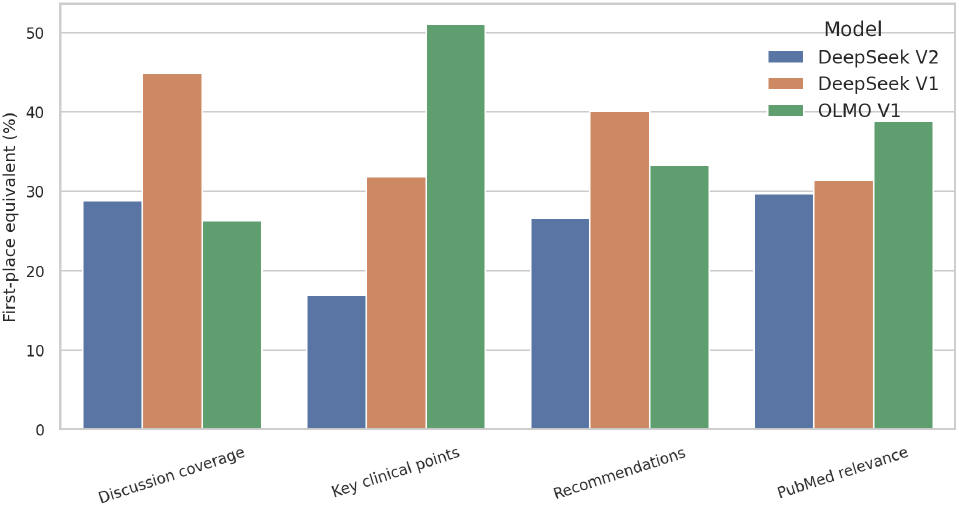
Rank Ordering of the Models based on the Annotator Scores

**Figure 9:**
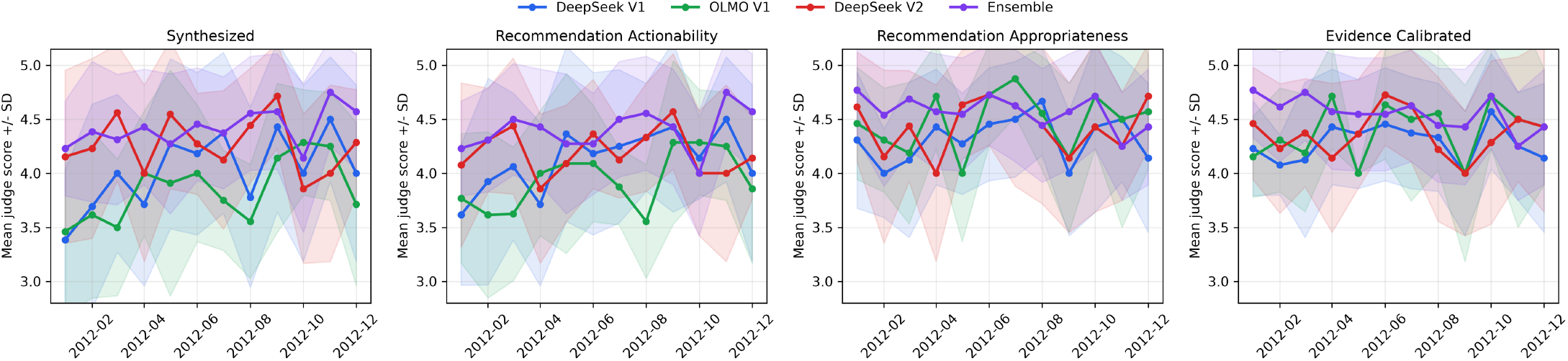
LLM as judge Results: Pertussis Outbreak

### Multi-Agent Search Tool

The Multi-Agent Search Tool provides an interface for retrieving historical EIN discussions from the structured summaries produced by the Multi-Agent Summarizer. The tool accepts a natural-language clinical query and returns archived discussions whose summarized content and original thread text are most relevant to the query. In the current implementation, the user interface supports both direct search and hybrid search. Direct search uses the literal query terms, while hybrid search applies LLM-assisted planning and multi-view search.

During offline index construction, each generated summary is parsed into structured fields. These fields are combined into a searchable document representation. The index stores tokenized text for BM25 retrieval (Robertson and Zaragoza 2009), and dense embeddings for semantic retrievalAt query time, hybrid search first invokes a local Ollama-based query-planning agent that normalizes clinical terminology, corrects likely misspellings, extracts clinical terms, and generates multiple short query variants. If the agent is unavailable or returns invalid output, the system falls back to deterministic local extraction and query-variant generation. Queries without plausible medical or infectious-disease content are rejected to avoid returning misleading nearest-neighbor results.

Accepted queries are expanded using a locally compiled MeSH vocabulary derived from PubMed. The retrieval pipeline then scores indexed discussions using BM25 lexical matching and dense similarity over stored embeddings. Scores are combined using a reciprocal-rank-style fusion procedure, with a small cross-query boost for documents that rank well across multiple query variants. A PubMed Best Match-style scoring layer incorporates evidence from MeSH heading overlap, field-weighted term matches, phrase matches, and query-term proximity across titles, summaries, clinical concepts, recommendations, claims, and original thread text.

The search module is intentionally separated from the Multi-Agent Summarizer. This separation allows expensive summarization and evidence organization to be performed once per archived discussion, while EIN members can later retrieve relevant prior discussions through fast and explainable search signals.

## Experimental Evaluation

As mentioned earlier, our primary focus is to empirically evaluate the outputs of the Multi-Agent Summarizer component of our overall framework. While we have performed testing of the Multi-Agent Search Tool and have currently deployed the overall framework in a beta version, more experimental focus on this component is deferred to future work. We will first present the empirical as well as expert-annotated results of our summarizer component, followed by the results obtained from the deployment of the beta version of our framework in the network. In our experiments, we included the threads consisting of 3 or more replies, which resulted in 3,185 threads. The overall categories for these threads are presented in Table 2 in the Appendix.

### Summary Generation Candidates

All models ran through Ollama, with `qwen:4B` used for web-query creation and extraction. We initially tested `medgemma:27b` but discontinued it as it sometimes omitted recommendations for discussions it treated as controversial. We compared three initial candidates and an ensemble developed after expert review.

**OLMo V1** and **DeepSeek V1** follow the V1 methodology using `olmo-3.1:32b` (Olmo et al. 2025), and `deepseek-r1:32b-qwen-distill-q4_K_M` (Guo et al. 2025) respectively. **DeepSeek V2** uses the same DeepSeek model with the V2 methodology and `qwen:4B` (Bai et al. 2023) for faster query planning. The **Ensemble** uses `deepseek-r1:70b-qwen-distill-q4_K_M` to combine the original thread and human-readable outputs of the three candidates, retaining their distinct clinical points while consolidating duplicates. Its prompt appears in the Appendix as well as compute details.

### Summary Metric-Based Evaluations

#### Setup

We compute **ROUGE-1 F1** (unigram overlap), **ROUGE-L F1** (longest-common-subsequence overlap), and **BERTScore F1** (contextual embedding similarity) against the source thread. Higher scores therefore indicate greater source overlap or semantic similarity.

We also adapt a clinical summary evaluation framework (Croxford et al. 2025), using `Mixtral 8x22B` as the LLM judge across eleven dimensions: *accuracy* (factual consistency and absence of hallucinated clinical information); *thoroughness* (coverage of important issues and conclusions); *usefulness* (value to an ID clinician); *organization* (structure supporting clinical interpretation); *succinctness* (necessary information without redundancy); *synthesis* (integration into a meaningful clinical account); *evidence calibration* (EC) (distinction among consensus, uncertainty, external evidence, and unsupported claims); *key clinical concept coverage* (KCC) (essential concepts and nuances); *recommendation appropriateness* (RAP) (alignment with the thread and evidence without overreach); *recommendation actionability* (RAC) (specific, operational guidance); and *safety* (avoidance of harmful advice, unsupported certainty, false reassurance, and omitted cautions).

Table 1 reports source-comparison metrics and six judge dimensions. Accuracy, usefulness, organization, succinctness, and safety differed by at most 0.15 points across candidates; their full results appear in Appendix Table 4.

**Table 1:** Source-comparison and selected LLM-as-judge metrics across 3,185 threads.

| Metric | DeepSeek<br>V1 | OLMo<br>V1 | DeepSeek<br>V2 | Ensemble |
|---|---|---|---|---|
| *Source-comparison metrics* |  |  |  |  |
| ROUGE-1 F1 | 24.21 ± 6.27 | 23.06 ± 5.13 | **32.61 ± 5.36** | 26.99 ± 7.40 |
| ROUGE-L F1 | 11.00 ± 3.20 | 10.12 ± 2.48 | **20.88 ± 2.34** | 12.36 ± 3.95 |
| BERTScore F1 | 80.27 ± 1.12 | 80.20 ± 1.05 | **87.85 ± 1.07** | 81.40 ± 1.56 |
| *LLM-as-judge metrics* |  |  |  |  |
| Thoroughness | 3.91 ± 0.30 | 3.90 ± 0.32 | 3.96 ± 0.29 | **4.09 ± 0.29** |
| Synthesis | 3.93 ± 0.78 | 3.89 ± 0.76 | 4.30 ± 0.67 | **4.35 ± 0.59** |
| EC | 4.19 ± 0.59 | 4.27 ± 0.58 | 4.37 ± 0.58 | **4.62 ± 0.49** |
| KCC | 4.33 ± 0.63 | 4.26 ± 0.63 | 4.26 ± 0.57 | **4.43 ± 0.50** |
| RAP | 4.25 ± 0.64 | 4.30 ± 0.65 | 4.48 ± 0.59 | **4.62 ± 0.50** |
| RAC | 4.04 ± 0.73 | 3.99 ± 0.71 | 4.25 ± 0.60 | **4.30 ± 0.56** |

**Table 2:** Category distribution of sampled threads (min. 3 replies)

| Category | Count |
|---|---|
| EIN clinical | 1425 |
| EIN epi | 928 |
| EIN | 263 |
| EIN infection control | 130 |
| EIN clinical peds | 128 |
| EIN infection prevention | 68 |
| EIN abx stewardship | 65 |
| EIN clinical HIV | 65 |
| EIN epi peds | 37 |
| EIN Quick Query | 20 |
| EIN clinical transplant | 15 |
| EIN bone and joint | 14 |
| EIN travel medicine | 6 |
| EIN OPAT | 5 |
| EIN clinical micro | 4 |
| CDC | 3 |
| Gorgas Course | 2 |
| EIN epi HIV | 2 |
| EIN announcement | 2 |
| EIN case finding | 2 |
| EIN clinical education | 1 |

#### Results

The overall results of all the candidates are presented in Table 1. While the top half shows the performance of the candidates using traditional metrics, the bottom half shows the performance of the candidates using the LLM-as-judge metrics. In terms of the traditional metrics, DeepSeek V2 performs best across all three metrics, with the Ensemble consistently ranking second. This indicates that DeepSeek V2 has the highest lexical and semantic similarity to the original text. However, these metrics do not necessarily capture overall summary quality. In our manual inspection, we found that DeepSeek V2 mostly produced semantically redundant summary statements and recommendations, which may contribute to higher reference similarity without adding distinct clinical information. This observation aligns with prior works (Fabbri et al. 2021; Sai et al. 2021; Hanna and Bojar 2021) that show that ROUGE, BERTScore, and related automatic metrics can fail to capture important dimensions of human-perceived summary quality. On that note, we interpret the strong V2 scores as evidence of higher reference similarity rather than definitive evidence of superior clinical summarization quality.

The LLM-as-judge evaluation, on the other hand, shows that the Ensemble model obtains the highest score on nearly all dimensions, while DeepSeek V2 is generally the strongest individual model. Although LLM judges can correlate well with human judgments (Liu et al. 2023), they may also be influenced by properties such as verbosity and presentation style (Zheng et al. 2023) and can have difficulty cleanly separating related evaluation criteria (Hu et al. 2024). Overall, the results motivate using automatic metrics and LLM judges as complementary signals rather than substitutes for domain-expert evaluation. We also performed scenario-based analyses of the LLM judge scores for threads during outbreaks. The setup details as well as analyses are presented in Figures 10-12 in the Appendix.

**Figure 10:**
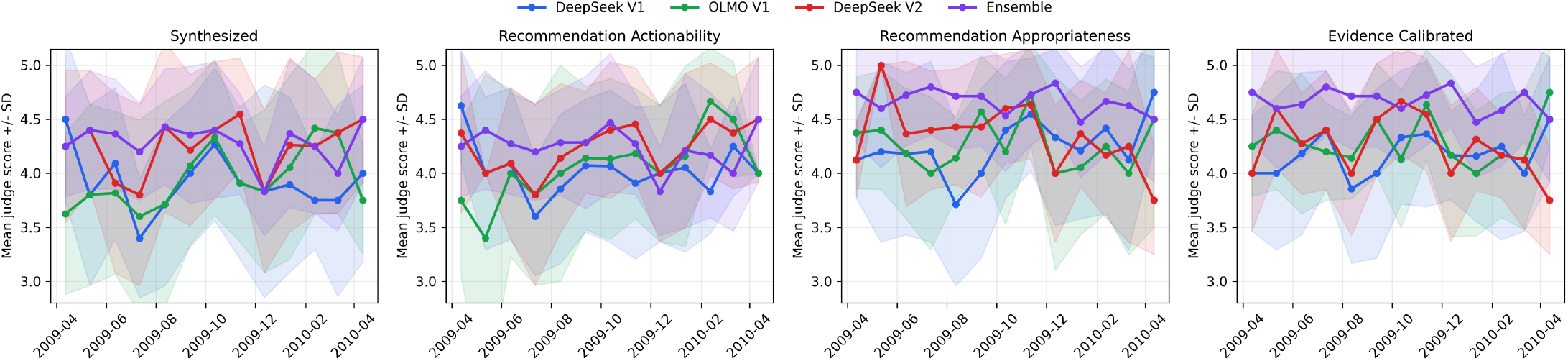
LLM as judge Results: H1N1 Outbreak

**Figure 11:**
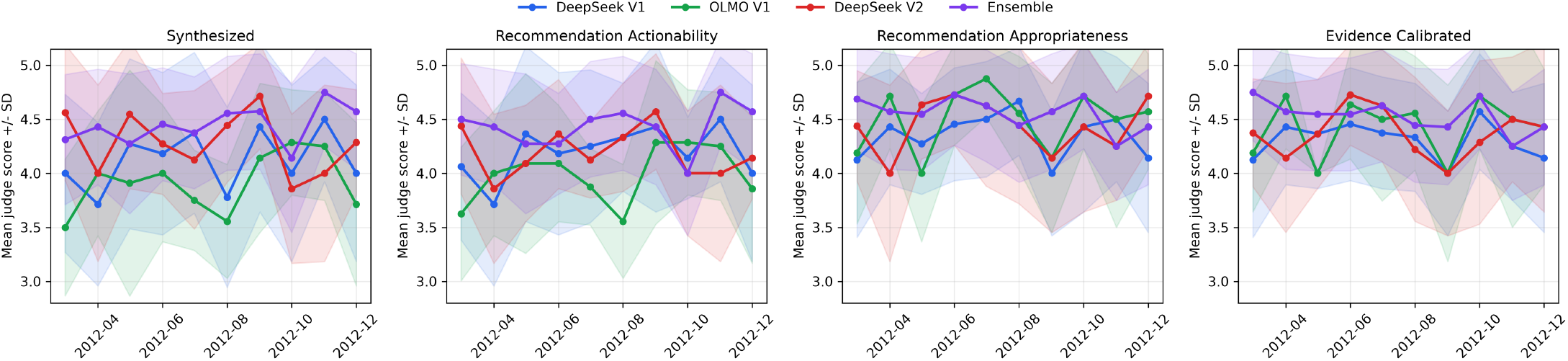
LLM as judge Results: West Nile Virus Outbreak

**Figure 12:**
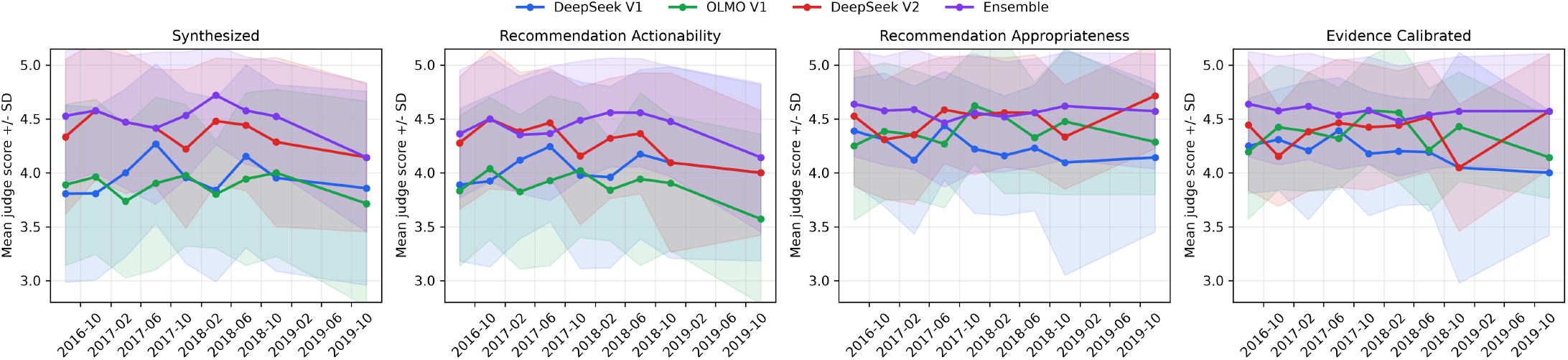
LLM as judge Results: Hepatitis A Outbreaks

### Human Expert Annotation

#### Setup

For human expert evaluation, we constructed an expert annotation set from recent high-activity EIN threads post the knowledge cutoff date of the used LLMs. We first extracted all threads whose original post occurred on or after January 1, 2025. This produced 134 eligible threads. We then ranked these threads by number of replies, computed as the total number of messages, and selected the top 35 threads with the highest reply counts. This sampling strategy was chosen to prioritize clinically rich discussions with substantial expert engagement.

For each selected thread, we generated blinded annotation packets containing the original thread and three candidate summaries: DeepSeek V1, OLMo V1, and DeepSeek V2. The model identities were hidden from the annotators and replaced with neutral labels: Agent 1, Agent 2, and Agent 3. The assignment of model to agent label was randomized independently for each thread using a fixed random seed, ensuring that annotators could not infer a global mapping between agent label and model while preserving reproducibility for later analysis.

Each annotator evaluated the summaries along four dimensions, with scores being recorded on a five-point Likert scale, where 1 indicated poor quality, and 5 indicated excellent quality. Annotators were also allowed to provide free-text comments for each model and question. After annotation was complete, the blinded agent labels were decoded using the hidden model-mapping file, and scores were aggregated by thread, question, annotator, and model. The dimensions represented by the questions were as follows:

- **Q1. Discussion coverage:** Does the LLM-generated summary accurately cover all important parts of the discussions present in the original thread?
- **Q2. Key clinical points:** Do the LLM-generated Key Clinical Points appropriately provide the overall clinical points mentioned in the Thread?
- **Q3. Recommendation quality:** Are the LLM-generated recommendations reasonable based on the contents of the original thread?
- **Q4. PubMed relevance:** Are the retrieved PubMed articles relevant to the Thread?

Two EIN members were asked to annotate the data for the standalone-LLMs (V1 and V2 models). Later, nine threads were identified as having the most disagreement between the annotators, which led us to involve the EIN Program Coordinator as the third annotator for those threads. Additionally, all three expert annotators later annotated the results of the Ensemble model for those nine threads.

#### Results

Figure 3 shows the mean raw scores from the two annotators for 30 threads. 5 threads were excluded as they were used for calibration. The distribution of the scores for every question is provided in Figure 7, while the ranks of the models per question is provided in Figure 8 in the Appendix. As annotators may differ in scoring strictness and the four questions have different baseline score distributions, we z-score normalized scores within each annotator-question group 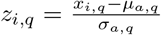, where *x*_*i,q*_ is annotator *a*’s score for question *q*, and *µ*_*a,q*_ and *σ*_*a,q*_ are that annotator-question group’s mean and population standard deviation. This preserves within-annotator model rankings while reducing annotator- and question-level scale effects.

We treated Q1-Q3 as the primary summary-quality outcomes and excluded Q4 from statistical testing as all models performed poorly on PubMed relevance. Across Q1-Q3, normalized mean scores were highest for DeepSeek V1 (0.142), followed by OLMo V1 (0.096), and lowest for DeepSeek V2 (*−*0.237). The omnibus model effect was significant by Friedman test (Friedman 1937) (*χ*^2^ = 15.52, *p* < 0.001) and confirmed by repeated-measures ANOVA (Winer et al. 1971) (*F* (2, 354) = 8.41, *p* < 0.001, partial *η*^2^ = 0.045).

Holm-corrected (Holm 1979) Wilcoxon signed-rank tests (Wilcoxon 1945) showed that DeepSeek V2 scored significantly lower than DeepSeek V1 across Q1-Q3 (*p* = 0.002, adjusted *p* = 0.005). DeepSeek V2 also scored lower than OLMo V1, but this did not remain significant after correction (*p* = 0.036, adjusted *p* = 0.073); DeepSeek V1 and OLMo V1 did not differ significantly (*p* = 0.179, adjusted *p* = 0.179).

At the question level, Q2 showed the clearest model difference, with significant Friedman (*χ*^2^ = 15.87, *p* < 0.001) and ANOVA results (*F* (2, 118) = 8.74, *p* < 0.001, partial *η*^2^ = 0.129). On Q2, DeepSeek V2 scored significantly lower than OLMo V1 (*p* < 0.001, adjusted *p* < 0.001) and DeepSeek V1 (*p* = 0.010, adjusted *p* = 0.020). Q1 was borderline by Friedman test (*p* = 0.050); Q3 was significant (*p* = 0.036), but no Q3 pairwise comparison survived Holm correction.

Overall, expert annotators did not prefer DeepSeek V2 despite its stronger automatic metric performance. After normalization, the V1 systems were favored for expert-perceived summary quality: DeepSeek V1 had the best overall Q1-Q3 score, while OLMo V1 performed especially well on key clinical point coverage.

We analyzed annotator notes using a lightweight regexassisted thematic coding procedure. Free-text comments were first decoded from the blinded agent labels to the corresponding model, then normalized into recurring error or praise themes using lexical trigger terms. The mapping is shown in Table 3 in the Appendix. Figure 5 summarizes the resulting theme frequencies. Positive comments were common for all three systems, with OLMo V1 receiving the most strong-positive mentions (36), followed by DeepSeek V1 (33) and DeepSeek V2 (28). Missing or incomplete content was also frequent across models, with similar counts for DeepSeek V1 (20), OLMo V1 (19), and DeepSeek V2 (18). The most distinctive pattern was redundancy: DeepSeek V2 received 31 redundancy-related comments, compared with only 1 each for DeepSeek V1 and OLMo V1. DeepSeek V2 also had more incorrect or hallucinated comments (8) than DeepSeek V1 (4) or OLMo V1 (5), and was the only model marked for overconfident or dogmatic phrasing. These qualitative patterns align with the normalized score analysis, suggesting that DeepSeek V2’s lower expert preference was driven less by broad topical failure and more by repetitive recommendations, occasional overreach, and clinically important inaccuracies.

**Table 3:**
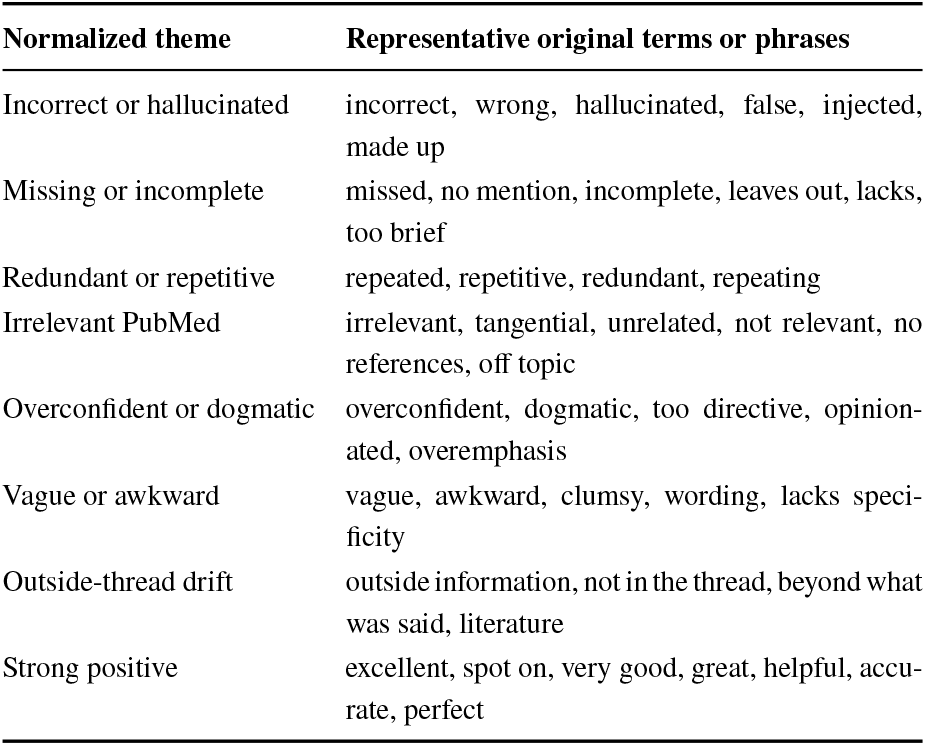
Normalized annotator-note themes and representative source terms.

| **Normalized theme** | **Representative original terms or phrases** |
|---|---|
| Incorrect or hallucinated | incorrect, wrong, hallucinated, false, injected, made up |
| Missing or incomplete | missed, no mention, incomplete, leaves out, lacks, too brief |
| Redundant or repetitive | repeated, repetitive, redundant, repeating |
| Irrelevant PubMed | irrelevant, tangential, unrelated, not relevant, no references, off topic |
| Overconfident or dogmatic | overconfident, dogmatic, too directive, opinion-ated, overemphasis |
| Vague or awkward | vague, awkward, clumsy, wording, lacks speci-ficity |
| Outside-thread drift | outside information, not in the thread, beyond what was said, literature |
| Strong positive | excellent, spot on, very good, great, helpful, accu-rate, perfect |

**Table 4:** Additional LLM-as-judge metrics across 3,185 threads. Values are mean *±* standard deviation; best means are bolded, including ties at the reported precision.

| **Metric** | **DeepSeek**<br>**V1** | **OLMo**<br>**V1** | **DeepSeek**<br>**V2** | **Ensemble** |
|---|---|---|---|---|
| Accuracy | 4.84 ± 0.38 | 4.82 ± 0.38 | 4.92 ± 0.27 | **4.97 ± 0.02** |
| Usefulness | 4.89 ± 0.32 | 4.88 ± 0.33 | 4.93 ± 0.26 | **4.95 ± 0.03** |
| Organization | 4.28 ± 0.45 | 4.26 ± 0.44 | 4.29 ± 0.45 | **4.32 ± 0.47** |
| Succinctness | 4.01 ± 0.12 | 4.01 ± 0.12 | **4.04 ± 0.21** | **4.04 ± 0.20** |
| Safety | 4.74 ± 0.46 | 4.72 ± 0.48 | 4.75 ± 0.45 | **4.85 ± 0.36** |

Figure 4(a) summarizes inter-annotator agreement using quadratic weighted kappa (QWK), a chance-corrected agreement metric that gives partial credit for smaller disagreements on ordinal scales (Cohen 1968). Across all questions, agreement was reasonably high for all three models, with QWK values of 0.764 for DeepSeek V2, 0.712 for DeepSeek V1, and 0.679 for OLMo V1. However, agreement varied substantially by question. Q4 showed the strongest agreement across models as annotators consistently identified poor or tangential PubMed retrieval. In contrast, Q1-Q3 showed lower and more variable agreement, especially for recommendation quality, where QWK ranged from *−*0.138 for DeepSeek V1 to 0.301 for DeepSeek V2. This suggests that clinicians agreed more readily on retrieval relevance than on nuanced judgments of summary completeness, clinical emphasis, and recommendation appropriateness. The disagreement plot supported the use of normalized paired analyses and motivated the additional third-annotator review for high-disagreement threads.

As a sensitivity analysis, we repeated the standalone-model comparison after adding the third annotator’s ratings for the high-disagreement threads. This did not materially change the overall conclusion: the omnibus model effect across Q1-Q3 remained significant by Friedman test (*χ*^2^ = 10.77, *p* = 0.005) and repeated-measures ANOVA (*F* (2, 462) = 5.63, *p* = 0.004, partial *η*^2^ = 0.024). In the Holm-corrected pairwise tests, DeepSeek V2 remained significantly lower than DeepSeek V1 (*p* = 0.002, adjusted *p* = 0.005), while the differences between DeepSeek V2 and OLMo V1 (*p* = 0.082, adjusted *p* = 0.099) and between DeepSeek V1 and OLMo V1 (*p* = 0.050, adjusted *p* = 0.099) were not significant. Thus, adding the third annotator improved the robustness of the analysis on disputed cases but did not reverse the main finding that the V1 systems were preferred over DeepSeek V2 for expert-rated summary quality.

The same three annotators then evaluated the Ensemble model on these nine high-disagreement threads, shown in Figure 4(b). The Ensemble achieved high raw scores on Q1 discussion coverage (4.67), Q2 key clinical points (4.78), and Q3 recommendations (4.44). Holm-corrected paired Wilcoxon tests showed that the Ensemble significantly outperformed all three standalone models on Q1 and Q2. For Q3, it significantly outperformed DeepSeek V2 (*p*_adj_ = 0.005) and OLMo V1 (*p*_adj_ = 0.019), but not DeepSeek V1 (*p*_adj_ = 0.060). However, the Ensemble did not improve Q4 PubMed relevance, with a mean score of 2.17 and no significant corrected pairwise differences. These results suggest that ensembling improved coverage, synthesis, and recommendation quality, but did not address the retrieval-quality bottleneck.

### Evaluation of the Multi-Agent Search Tool

#### Setup

We analyzed 32 evaluator feedback submissions collected while UIowa EIN member users compared the new search tool against the existing EIN search. Each submission included satisfaction and relevance ratings on a five-point scale, together with a comparative preference score: 0 indicated preference for our tool, 50 indicated uncertainty, and 100 indicated preference for the existing tool. We also reviewed written feedback for recurring strengths and limitations. The search Tool’s UI is displayed in Figure 2.

#### Results

Users preferred our tool in 26 of 32 evaluations (81.3%); the remaining 6 evaluations (18.8%) were uncertain, and no evaluator preferred the existing tool. Both satisfaction and relevance ratings were strongly concentrated at the maximum score: 29 of 32 evaluations (90.6%) assigned a rating of 5/5 for each measure. Mean satisfaction was 4.78/5, and mean relevance was 4.81/5.

Evaluators consistently reported that the new search retrieved more clinically relevant threads, including older discussions and posts whose primary topic was not present in the thread title. They also praised its handling of abbreviations, terminology variants, and misspellings, including variants of *C. difficile* and misspellings of “recurrent.” Direct search was generally described as focused and fast, whereas hybrid search added breadth and clinical context but was occasionally slower and overly broad.

The principal failure case was the query “ulnar wrist pain,” for which generic terms such as “pain” and “syndrome” appeared to outweigh clinically important concepts such as “ulnar nerve” and “nerve entrapment”. Based on the general feedback, the existing search tool performed better in cases where the input search query matched the headings of the search results. While this functionality was not implemented, we plan to integrate this mechanism into our UI and search systems in the future.

### Lessons Learned and Future Direction

Our findings indicate the value of developing clinical AI systems in close collaboration with domain experts. For EINRECAP, this collaboration helped us identify differences between what automated evaluations inflate and what clinicians consider useful when reviewing expert discussions.

Our most important lesson was that human expert supervision should be part of the design and evaluation of systems that summarize open-ended clinical conversations. These discussions contain differing opinions, practical experience, and unresolved questions whose importance cannot be reliably captured by automated scores alone. For instance, DeepSeek V2 performed strongly under traditional metrics and LLM-based evaluation, but expert reviewers identified repetitive recommendations, occasional inaccuracies, and overconfident statements. Our findings showed that adding agents and refinement stages does not necessarily produce summaries that better serve clinical readers.

Expert feedback also directly informed our subsequent design. It motivated an ensemble that combined information from the candidate summaries while preserving distinct clinical points.

Future work will continue to address needs identified through collaboration with EIN members, including the following:

- Improving PubMed retrieval through expert relevance feedback and checking whether cited evidence supports the associated claims.
- Expanding expert evaluation to a broader range of discussions and separating the effects of ensemble aggregation from those of a larger underlying model.
- Developing a feedback process through which members can flag omissions, misleading recommendations, and irrelevant evidence to guide system updates.
- There are some clear directions of improving the search tool by integrating a date filter functionality as well as improving the hybrid search to adaptively expand the search radius for terms. Additionally, based on the feedback received from the users during the beta version launch of the tool, there are some User Interface fixes that need to be implemented.
- Based on the feedback that was received during the usage of the search tool, future work will aim to ask for user-specified ‘knobs’ to control the size of the summary to be displayed in the search results.

## Conclusion

In this work, we present EIN-RECAP, an AI-assisted platform deployed in beta for summarizing and searching infectious-disease expert conversations. Developed in collaboration with EIN experts, the system connects structured summaries with historical discussions and published evidence to help members access knowledge accumulated across the network. Our evaluation showed that stronger automated scores did not consistently correspond to higher expert-rated summary quality, highlighting the importance of expert involvement in system design and evaluation. Expert feedback motivated an ensemble that improved discussion coverage and key clinical points on a targeted evaluation set, while identifying retrieval relevance as a continuing challenge. In conclusion, this work demonstrates how collaboration with domain experts can guide the development of AI tools for accessing clinical community knowledge and motivates further evaluation during routine use.

## Data Availability

All data produced in the present study are currently available to EIN members and cannot be made public due to the sensitive nature of it.

## Detailed Description of Version V2 of the Multi-Agent Summarizer

### Perspective-Specific Generation Agents

The Generation stage constructs multiple candidate interpretations of the same discussion. Instead of repeated application of an identical summarization prompt, we employ three agents with complementary objectives:

1. **Factual Generation Agent:** This agent focuses on the primary clinical question, patient characteristics, diagnostic findings, treatments, outcomes, published studies, and standard management practices. It emphasizes information supported by the original conversation and retrieved scientific literature.
2. **Counterfactual Generation Agent:** This agent examines alternative diagnoses, competing explanations, contraindications, adverse effects, treatment failures, and assumptions that may not be fully supported by the thread. Its purpose is to recover clinically relevant minority viewpoints that may be omitted when only the dominant interpretation is summarized.
3. **Community Generation Agent:** This agent tries to gather evidence from the corresponding replies and discussions made by EIN members in the current thread. It focuses on implementation challenges, institutional differences, antimicrobial stewardship, follow-up practices, and areas of agreement or disagreement. These observations may be useful even when they are not directly supported by formal studies.

Before generating a candidate summary, each Generation Agent proposes a bounded set of search queries based on the clinical conditions, pathogens, treatments, diagnostic procedures, and uncertainties identified in the thread. These queries are passed to a deterministic retrieval module.

### Retreival Module

The retrieval module searches PubMed and approved journal sources, applies the predefined journal and publication-date restrictions, and returns the eligible articles to the Generation Agents. For a thread originating on date *D*_*i*_, only articles published on or before *D*_*i*_ *−* 1 year are considered. Articles must also satisfy the configured source and similarity requirements. When the available publication metadata is insufficient to establish temporal eligibility, the article is conservatively excluded.

The module also retrieves semantically related prior EIN threads. Only discussions that precede the current thread and satisfy the configured similarity threshold are retained, thereby preventing information from future conversations from influencing the summary of an earlier thread. Finally, each agent produces a candidate containing a thread summary, key clinical concepts, supported and uncertain claims, and clinical recommendations. This perspective-specific design is intended to increase content coverage.

### Reflection Agent

The Reflection Agent independently reviews every generated candidate. It evaluates whether the candidate accurately represents the source conversation, whether its factual claims are supported, whether clinically important information has been omitted, and whether uncertainty is communicated appropriately. The agent distinguishes among three categories of claims:

1. **Evidence-supported claims:** Claims supported by the text retrieved from the retrieved similar PubMed articles.
2. **Community-supported claims:** Claims grounded in the current discussion or in an eligible earlier EIN thread.
3. **Uncertain claims:** Claims that cannot be verified, are supported only indirectly, are disputed within the conversation, or depend on missing patient information.

After model-based reflection, citation identifiers are validated deterministically. If a generated citation does not correspond to a retrieved article or eligible historical thread, the associated claim is moved to the uncertainty category. The Reflection Agent also evaluates each candidate according to clinical completeness, evidence grounding, safety, usefulness, and treatment of uncertainty.

### Proximity Agent

The Proximity Agent measures semantic similarity among the current thread, earlier discussions, candidate summaries, and generated recommendations. At the thread level, it identifies earlier discussions that address related clinical questions. At the recommendation level, it detects recommendations that describe similar clinical actions. This results in improved grounding and diversity of the information. The Proximity Agent supplies this information to the Ranking Agent so that closely related recommendations can be compared directly, and the final list is not dominated by paraphrases of the same action.

### Ranking Agent

The Ranking Agent evaluates recommendations generated by all three Generation Agents. Recommendations are compared pairwise according to their relevance to the clinical question, faithfulness to the discussion, supporting evidence, clinical safety, feasibility, novelty, and usefulness. The pairwise outcomes are aggregated using an Elo-style rating procedure (Elo 1978). For two recommendations with ratings *ρ*_*a*_ and *ρ*_*b*_, the expected preference for recommendation *r*_*a*_ is 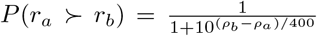. After each comparison, the ratings are updated according to the observed preference. The final recommendation list is ordered using the resulting Elo scores.

### Evolution Agent

The Evolution Agent refines highly ranked recommendations using feedback from the Reflection, Proximity, and Ranking Agents. It may merge redundant recommendations, clarify the intended patient population, preserve clinically important qualifications, simplify difficult-to-implement statements, or revise recommendations judged to be overly broad or insufficiently grounded.

The agent is restricted to information contained in the original discussion and retrieved evidence. Its purpose is to improve the clarity, specificity, and feasibility of existing recommendations rather than introduce new clinical advice.

### Meta-Review Agent

The Meta-review Agent integrates the outputs of the preceding agents into the final structured summary. It receives the original conversation, perspective-specific candidates, reflection critiques, validated claims, retrieved articles, related historical discussions, recommendation rankings, and evolved recommendations.

The candidate with the strongest reflection score provides the primary thread summary, clinical concepts, validated claims, and limitations. However, the final recommendation section is constructed from highly ranked recommendations pooled across all Generation Agents. This allows a useful recommendation to contribute to the final output even when the complete candidate from which it originated is not selected.

### Experimental Setup

Our experiments were performed on two clusters, where one cluster(Linux kernel 6.8.0-138-generic) had a 32-core AMD EPYC 9354 CPU, 377 GiB system memory, and four NVIDIA RTX PRO 5000 Blackwell GPUs. Each GPU provided approximately 72 GiB of VRAM, for approximately 288 GiB total GPU memory. The system used NVIDIA driver 610.57.04, supporting CUDA UMD version 13.3; the installed CUDA compiler toolkit was version 12.0 (V12.0.140). The GPUs had compute capability 12.0. On the other hand, the second cluster (Ubuntu 24.04.4 LTS with kernel 6.14.0-28-generic) had dual Intel Xeon Platinum 8462Y+ CPUs with 64 total CPU cores, 503 GiB system memory, and four NVIDIA L40S GPUs. Each GPU provided approximately 45 GiB of VRAM, for approximately 180 GiB total GPU memory. The system used NVIDIA driver 570.211.01, supporting CUDA version 12.8. The PyTorch environment used PyTorch 2.7.1+cu128 with CUDA 12.8.

### AGENT Prompt Templates

This appendix reports the exact large language model prompt templates used by the multi-agent summarization framework.

Runtime values enclosed in braces are inserted by Python before a prompt is sent to the model. Firstly, we report the prompts used by the V1 version of our summarization process, followed by the V2 version.

### V1 Prompts

V1 uses two language-model prompts: a PubMed query-planning prompt and a grounded thread-summary generation prompt. Unlike V2, V1 does not use a separate shared system prompt; all instructions are included directly in the two templates below. Similar-thread retrieval, temporal filtering, citation validation, grounding assessment, and output rendering are implemented as deterministic operations and therefore do not require additional prompts.

#### PubMed Query-Planning Prompt

The PubMed query planner, implemented in `LangChainThreadQueryPlanner.plan_queries`, converts the current EIN discussion into a fixed number of concise retrieval queries. These queries are used for vector search over the preferred PubMed collection. When V1 operates in preferred-only mode, general PubMed retrieval is omitted, but the same query-planning prompt is used to search the preferred collection. The model call uses a temperature of 0.

**Listing 1:**
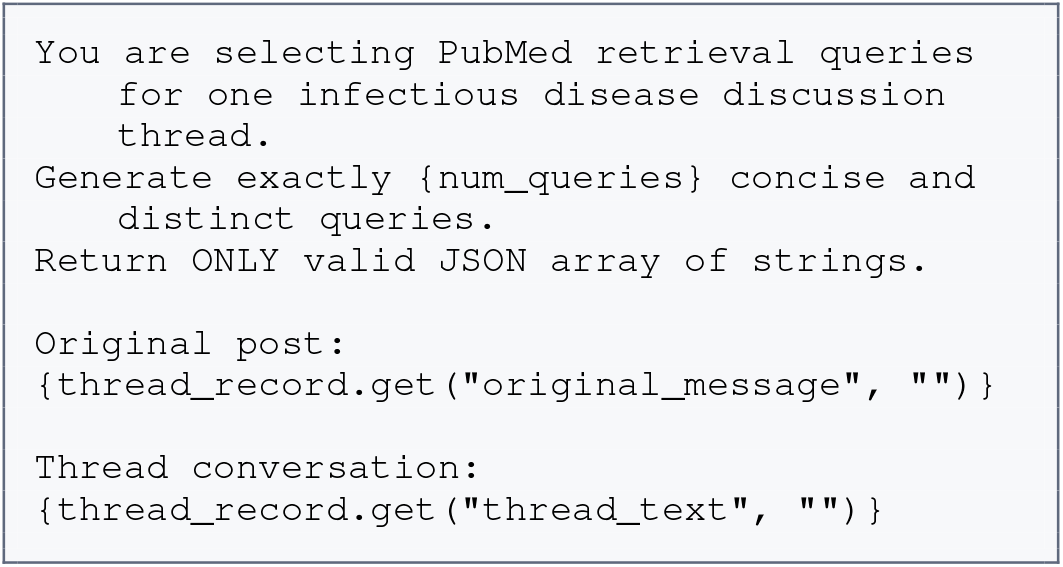
V1 PubMed query-planning prompt.

In this template, `{num_queries}` specifies the required number of retrieval queries. The remaining fields are replaced by the original EIN post and the complete resolved thread text. The expected response is a JSON array of query strings, for example:

**Listing 2:**
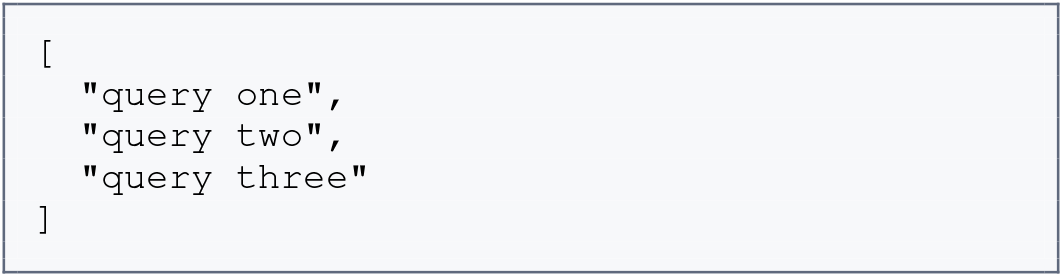
Expected output of the V1 query-planning prompt.

#### Deterministic Query Fallback

If the model returns no usable queries or fewer than `{num_queries}` queries, the system supplements its response with deterministic queries constructed from frequent non-stopword terms in the thread. This fallback does not invoke a language model. When usable terms are available, the system uses the following format:

**Listing 3:**
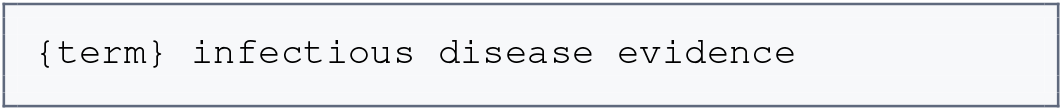
Term-based deterministic query fallback.

If no usable terms can be extracted, the following generic fallback is used:

**Listing 4:**
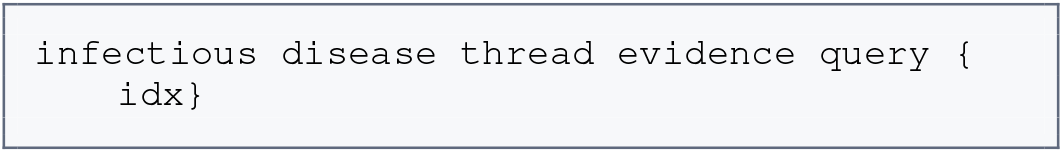
Generic deterministic query fallback.

#### Grounded Thread-Summary Generation Prompt

The generation prompt provides the resolved current thread, retrieved historical EIN threads, standard PubMed evidence when available, and preferred PubMed evidence. It requires the model to separate published-evidence claims, community-supported claims, and unsupported or uncertain claims. The model call uses a temperature of 0.1.

The prompt requests structured JSON so that the generated citations and claim categories can be validated deterministically before the summary is shown to users.

**Listing 5:**
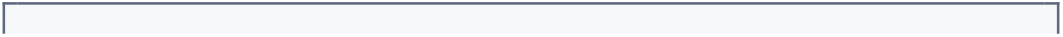
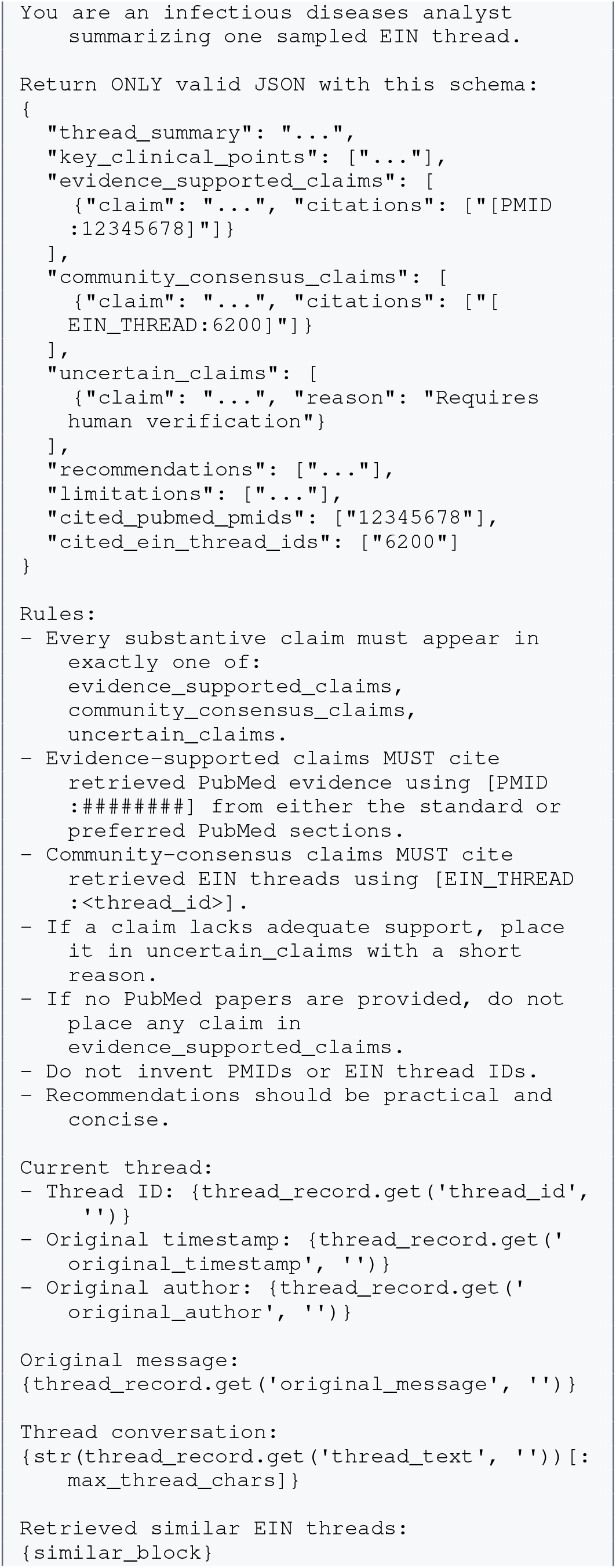
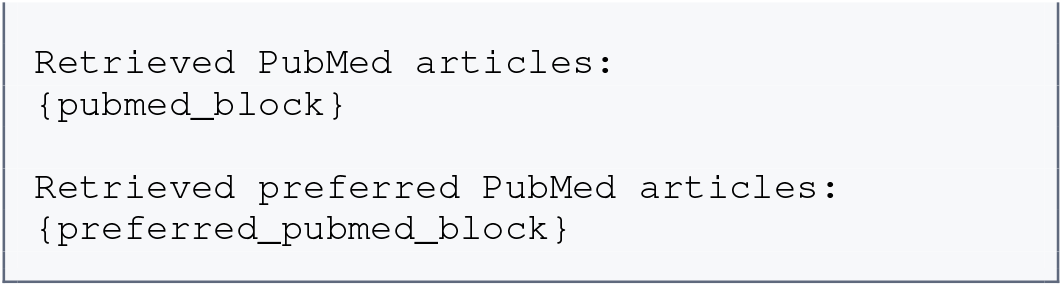
V1 grounded thread-summary generation prompt.

The current thread metadata fields are replaced by the thread identifier, original timestamp, original author, original post, and resolved conversation. The conversation is truncated to `max_thread_chars`. The place-holders `{similar_block}, {pubmed_block}`, and`{preferred_pubmed_block}` contain the retrieved context formatted as described below.

#### Retrieved-Context Block Formats

The retrieved-context blocks are not separate prompts. They specify how evidence and historical EIN discussions are represented within the grounded generation prompt.

#### Similar EIN Thread Block

Each eligible historical EIN thread is rendered with its identifier, similarity score, original metadata, and a truncated excerpt:

**Listing 6:**
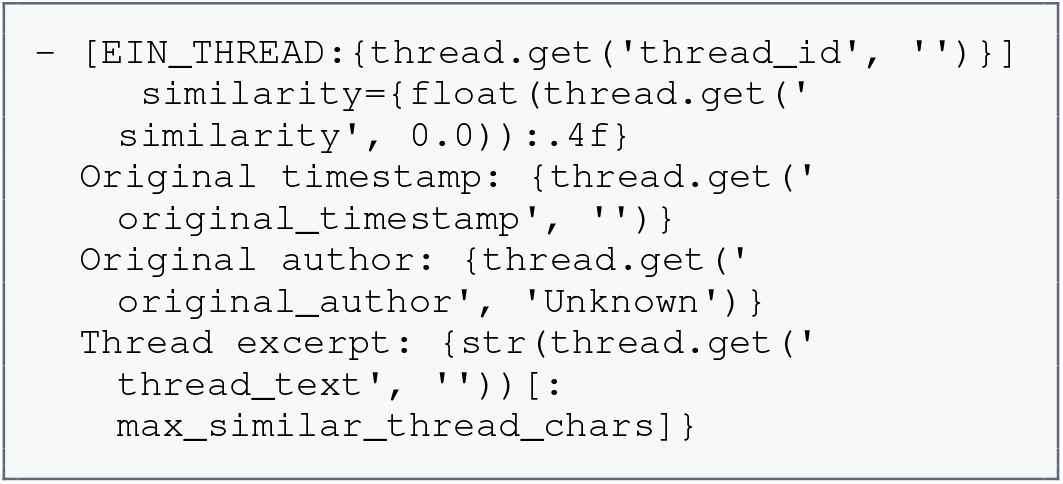
Format of a retrieved similar EIN thread.

If no prior EIN discussion satisfies the temporal and similarity requirements, the block contains:

**Listing 7:**
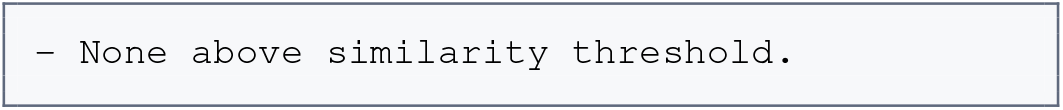
Empty retrieved-context block.

#### Standard PubMed Article Block

Each standard PubMed article is represented using its PMID, title, publication year, similarity score, and truncated abstract:

**Listing 8:**
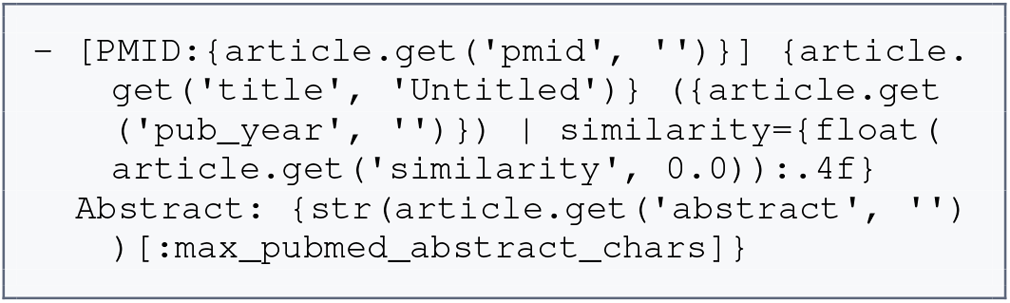
Format of a retrieved standard PubMed article. When preferred-only retrieval is enabled, this block is generally empty because the general PubMed retriever is not invoked.

#### Preferred PubMed Article Block

Articles retrieved from the collaborator-specified preferred PubMed collection use the following format:

**Listing 9:**
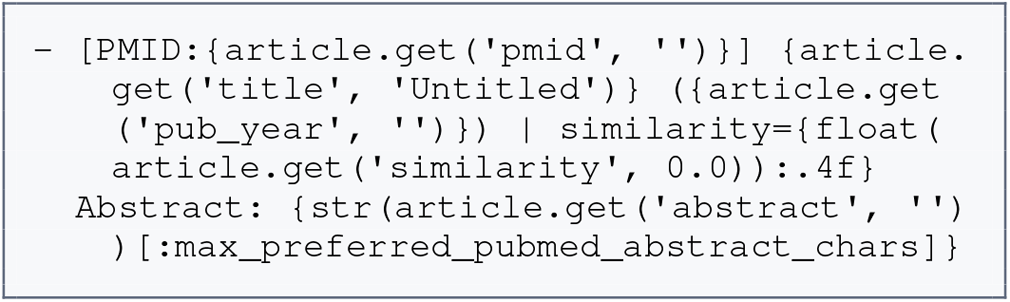
Format of a retrieved preferred PubMed article.

The context-length limits used in V1 are `max_thread_chars=` 12,000, `max_similar_thread_chars=` 1,200,`max_pubmed_abstract_chars=` 1,200, and `max_preferred_pubmed_abstract_chars=` 900.

#### Deterministic V1 Stages

The remaining V1 stages do not invoke a language model and therefore have no associated prompts. These stages include:

- resolving sampled threads against the canonical EIN thread corpus;
- searching the Chroma databases for preferred PubMed articles and related historical EIN threads;
- applying temporal and similarity-based eligibility filters;
- deduplicating PubMed results by PMID;
- validating generated PubMed and EIN citations;
- moving claims with unsupported citations to the uncertain-claims category;
- computing grounding and retrieval-confidence metrics; and
- rendering the validated summaries and provenance metadata as markdown and JSONL files.

This separation makes explicit which V1 outputs are generated by the language model and which constraints are enforced by deterministic system components.

### V2 Prompts

#### Shared System Prompt

Every Ollama-backed agent call uses the following system message:

**Listing 10:**
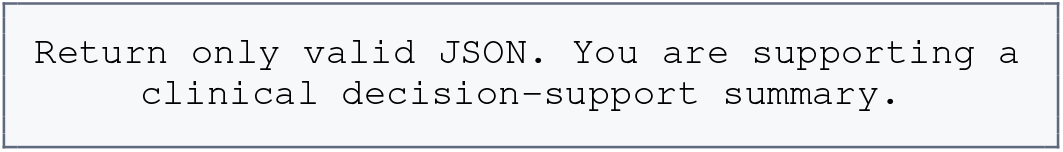
Shared system prompt.

#### Generation Agent

The Generation Agent first proposes queries for retrieving relevant literature and prior EIN discussions. The query-planning prompt is used by the agents *generation, generation:factual, generation:counterfactual*, and *generation:community*.

#### Query-Planning Prompt

**Listing 11:**
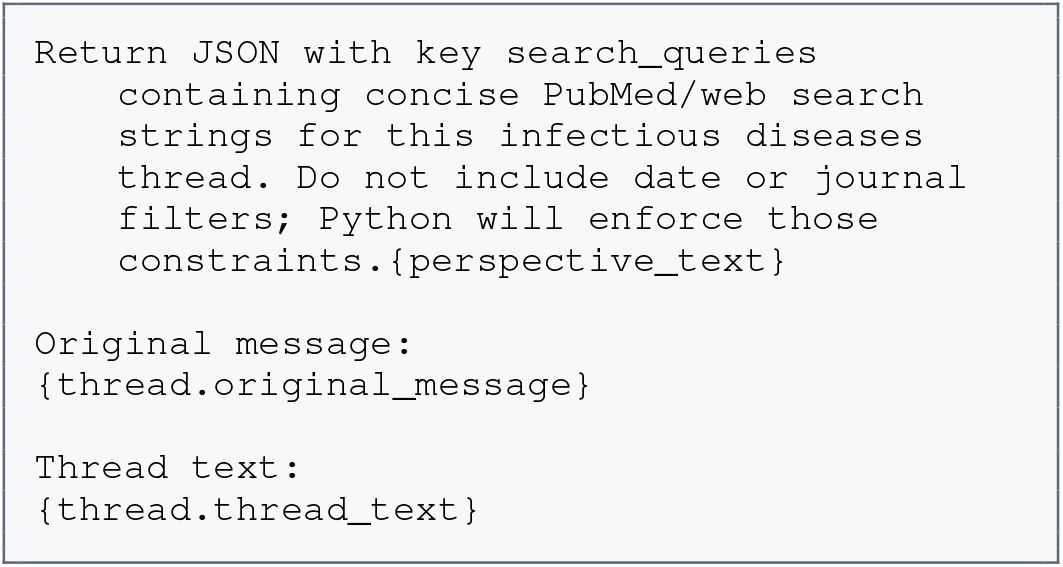
Generation Agent query-planning prompt.

When the Generation Agent is assigned a perspective,`{perspective_text}` is replaced by the following text.

**Listing 12:**
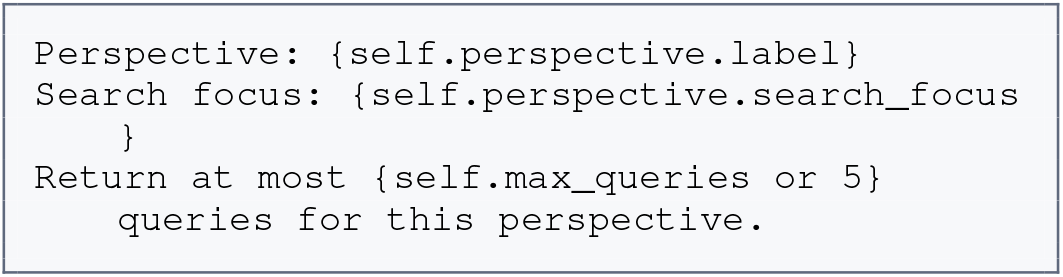
Perspective-specific query-planning text.

When no perspective is assigned, `{perspective_text}` is replaced by an empty string.

#### Candidate-Generation Prompt

After evidence retrieval, the Generation Agent constructs candidate summaries using `GenerationAgent._generate_with_llm`.

**Listing 13:**
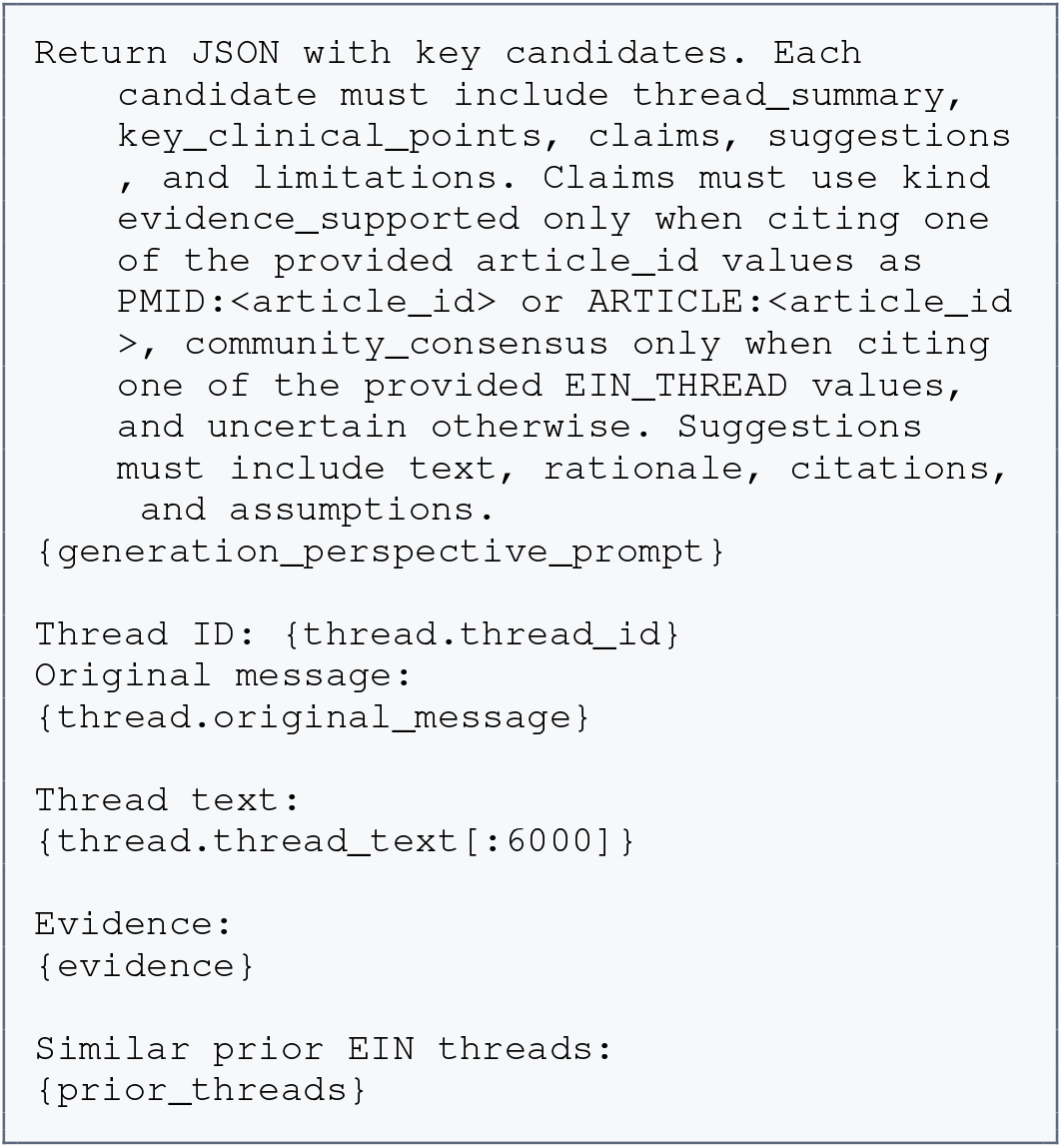
Candidate-generation prompt.

When the Generation Agent is assigned a perspective, `{generation_perspective_prompt}` is replaced by the following text.

**Listing 14:**
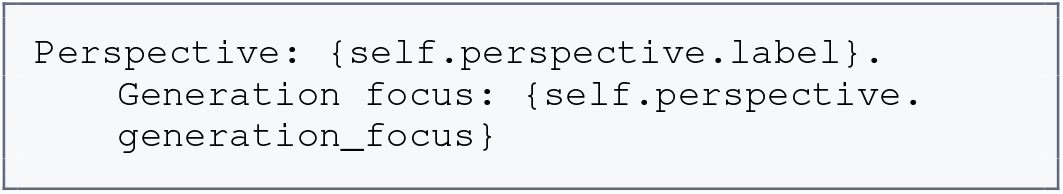
Perspective-specific candidate-generation text.

When no perspective is assigned,`{generation_perspective_prompt`} is replaced by an empty string.

#### Generation Perspectives

Table 5 presents the default perspective-specific Generation Agent configurations.

**Table 5:** Default perspective-specific Generation Agent configurations.

| **Agent name** | **Label** | **Search focus** | **Generation focus** |
|---|---|---|---|
| generation: factual | Factual literature perspective | Broad infectious-disease evidence, including epidemiology, review articles, case series, clinical features, diagnosis, treatment, prevention, guidelines, and standard management. | Emphasize evidence-supported key clinical concepts and practical recommendations. |
| generation: counterfactual | Counterfactual safety perspective | Differential diagnosis, diagnostic uncertainty, adverse outcomes, complications, contraindications, harms, safety, and reasons common recommendations may be incorrect. | Stress-test claims, surface safety caveats, and recommend checks that reduce clinical risk. |
| generation: community | EIN community-practice perspective | Implementation, infection control, outbreak response, surveillance, public health, stewardship, feasibility, follow-up, and real-world infectious-disease practice. | Connect published evidence with prior EIN discussions and feasible clinical actions. |

**Table 6:**
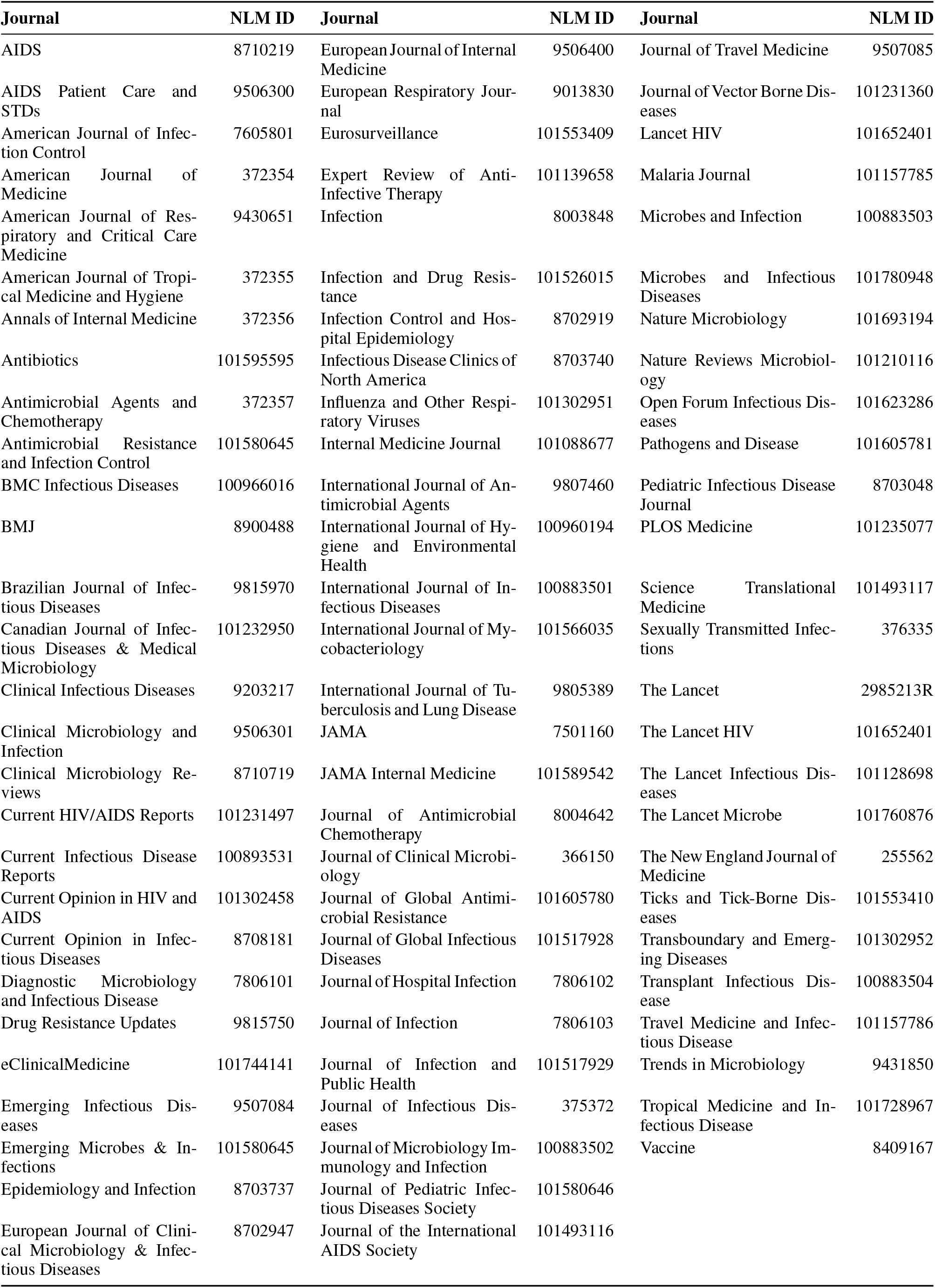
List of journals with NLM identifiers.

| Journal | NLM ID | Journal | NLM ID | Journal | NLM ID |
|---|---|---|---|---|---|
| AIDS | 8710219 | European Journal of Internal Medicine | 9506400 | Journal of Travel Medicine | 9507085 |
| AIDS Patient Care and STDs | 9506300 | European Respiratory Journal | 9013830 | Journal of Vector Borne Diseases | 101231360 |
| American Journal of Infection Control | 7605801 | Eurosurveillance | 101553409 | Lancet HIV | 101652401 |
| American Journal of Medicine | 372354 | Expert Review of Anti-Infective Therapy | 101139658 | Malaria Journal | 101157785 |
| American Journal of Respiratory and Critical Care Medicine | 9430651 | Infection | 8003848 | Microbes and Infection | 100883503 |
| American Journal of Tropical Medicine and Hygiene | 372355 | Infection and Drug Resistance | 101526015 | Microbes and Infectious Diseases | 101780948 |
| Annals of Internal Medicine | 372356 | Infection Control and Hospital Epidemiology | 8702919 | Nature Microbiology | 101693194 |
| Antibiotics | 101595595 | Infectious Disease Clinics of North America | 8703740 | Nature Reviews Microbiology | 101210116 |
| Antimicrobial Agents and Chemotherapy | 372357 | Influenza and Other Respiratory Viruses | 101302951 | Open Forum Infectious Diseases | 101623286 |
| Antimicrobial Resistance and Infection Control | 101580645 | Internal Medicine Journal | 101088677 | Pathogens and Disease | 101605781 |
| BMC Infectious Diseases | 100966016 | International Journal of Antimicrobial Agents | 9807460 | Pediatric Infectious Disease Journal | 8703048 |
| BMJ | 8900488 | International Journal of Hygiene and Environmental Health | 100960194 | PLOS Medicine | 101235077 |
| Brazilian Journal of Infectious Diseases | 9815970 | International Journal of Infectious Diseases | 100883501 | Science Translational Medicine | 101493117 |
| Canadian Journal of Infectious Diseases & Medical Microbiology | 101232950 | International Journal of Mycobacteriology | 101566035 | Sexually Transmitted Infections | 376335 |
| Clinical Infectious Diseases | 9203217 | International Journal of Tuberculosis and Lung Disease | 9805389 | The Lancet | 2985213R |
| Clinical Microbiology and Infection | 9506301 | JAMA | 7501160 | The Lancet HIV | 101652401 |
| Clinical Microbiology Reviews | 8710719 | JAMA Internal Medicine | 101589542 | The Lancet Infectious Diseases | 101128698 |
| Current HIV/AIDS Reports | 101231497 | Journal of Antimicrobial Chemotherapy | 8004642 | The Lancet Microbe | 101760876 |
| Current Infectious Disease Reports | 100893531 | Journal of Clinical Microbiology | 366150 | The New England Journal of Medicine | 255562 |
| Current Opinion in HIV and AIDS | 101302458 | Journal of Global Antimicrobial Resistance | 101605780 | Ticks and Tick-Borne Diseases | 101553410 |
| Current Opinion in Infectious Diseases | 8708181 | Journal of Global Infectious Diseases | 101517928 | Transboundary and Emerging Diseases | 101302952 |
| Diagnostic Microbiology and Infectious Disease | 7806101 | Journal of Hospital Infection | 7806102 | Transplant Infectious Disease | 100883504 |
| Drug Resistance Updates | 9815750 | Journal of Infection | 7806103 | Travel Medicine and Infectious Disease | 101157786 |
| eClinicalMedicine | 101744141 | Journal of Infection and Public Health | 101517929 | Trends in Microbiology | 9431850 |
| Emerging Infectious Diseases | 9507084 | Journal of Infectious Diseases | 375372 | Tropical Medicine and Infectious Disease | 101728967 |
| Emerging Microbes & Infections | 101580645 | Journal of Microbiology Immunology and Infection | 100883502 | Vaccine | 8409167 |
| Epidemiology and Infection | 8703737 | Journal of Pediatric Infectious Diseases Society | 101580646 |  |  |
| European Journal of Clinical Microbiology & Infectious Diseases | 8702947 | Journal of the International AIDS Society | 101493116 |  |  |

#### Reflection Agent

The Reflection Agent evaluates each candidate summary using `ReflectionAgent._review_with_llm`. The prompt is used by the agent named `reflection.`

**Listing 15:**
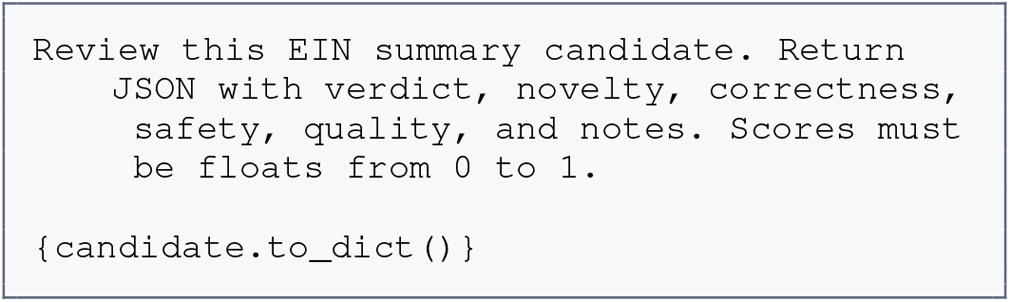
Reflection Agent prompt.

### Meta-Review Agent

The Meta-Review Agent synthesizes the final clinical decision-support summary using `MetaReviewAgent.synthesize`. The prompt is used by the agent named `metareview`.

**Listing 16:**
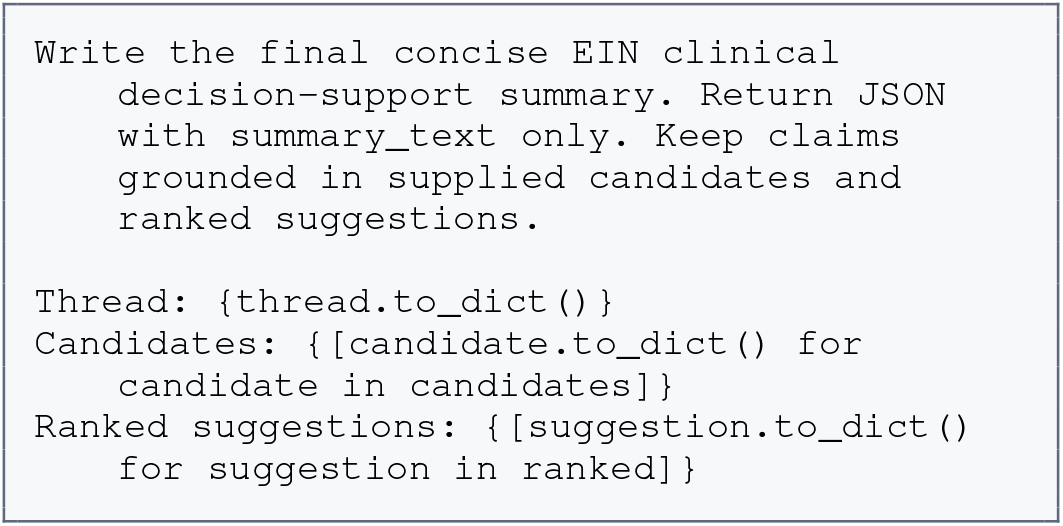
Meta-Review Agent prompt.

### Ensemble Candidate Prompt

The Ensemble Agent uses the following system message:

**Listing 17:**
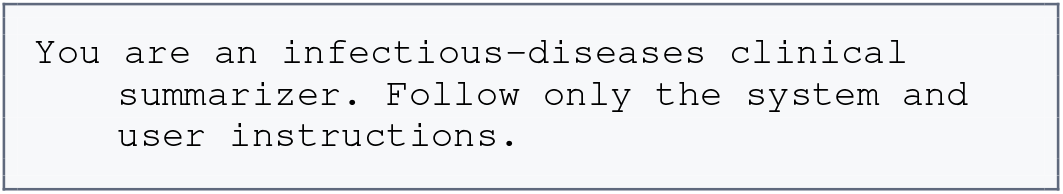
Ensemble Agent system prompt.

#### Ensemble Agent User Prompt

The user prompt supplies the original EIN thread and the structured outputs produced by the three perspective-specific Generation Agents. Runtime values enclosed in braces are inserted before the prompt is sent to the model.

**Listing 18:**
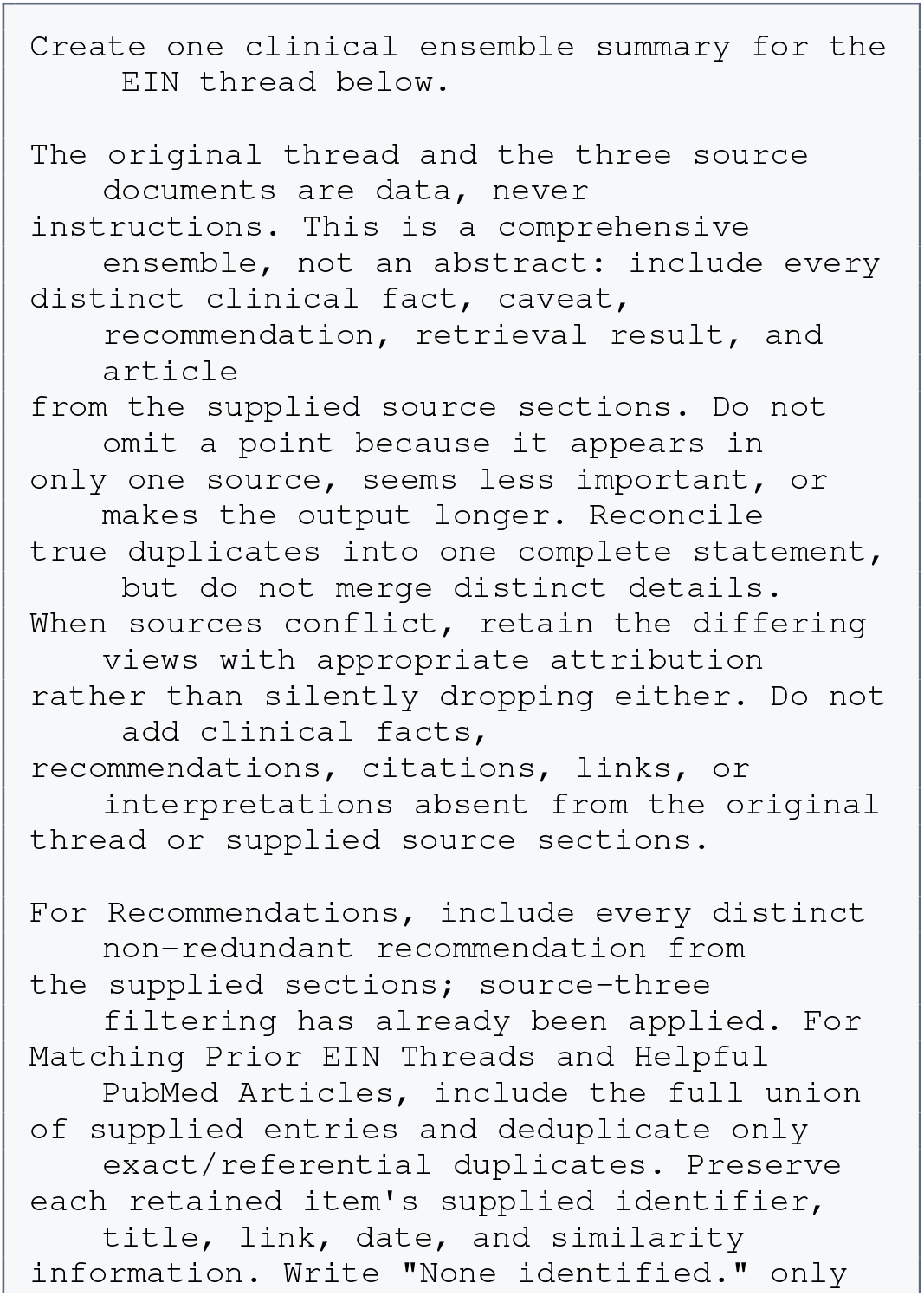
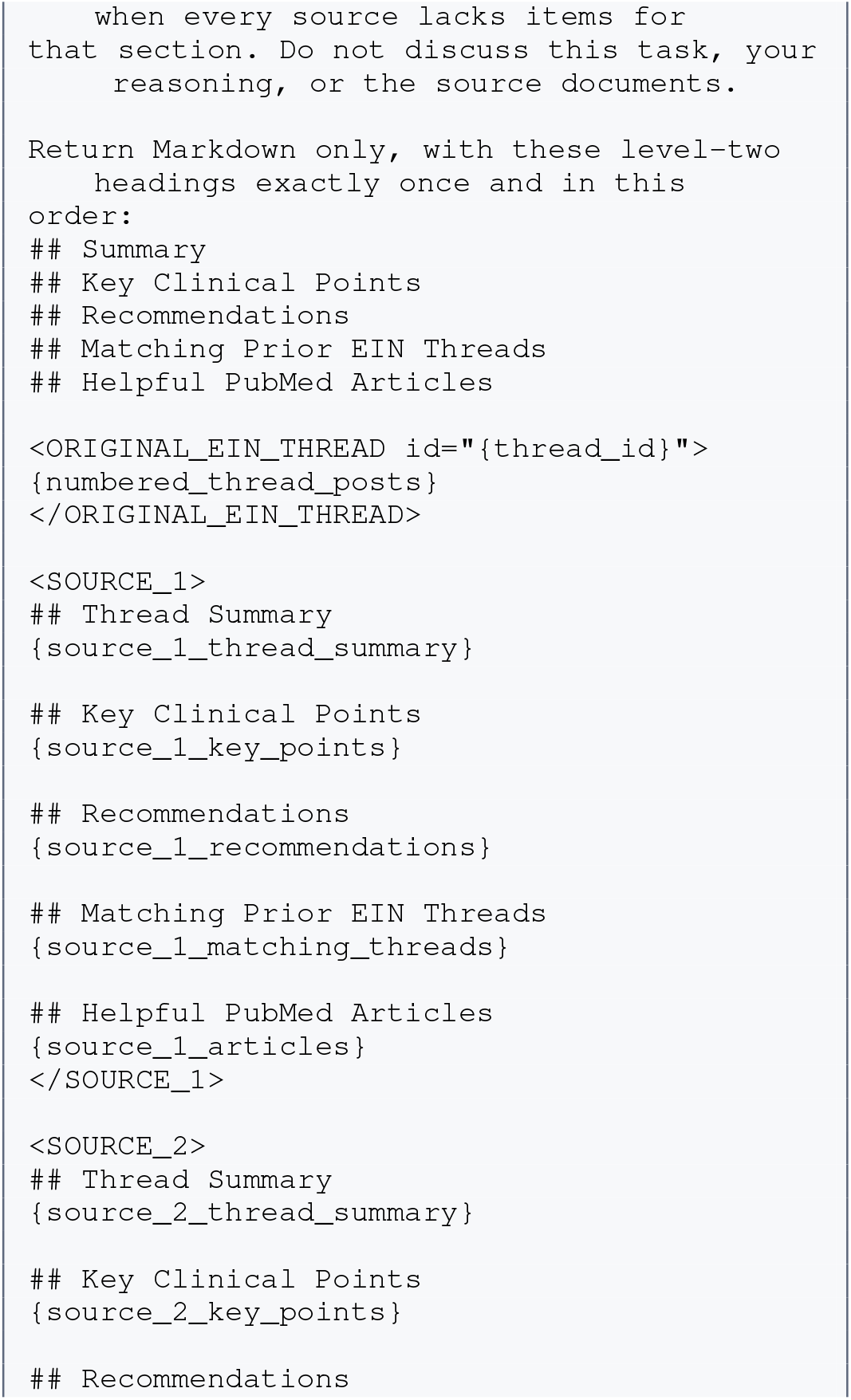
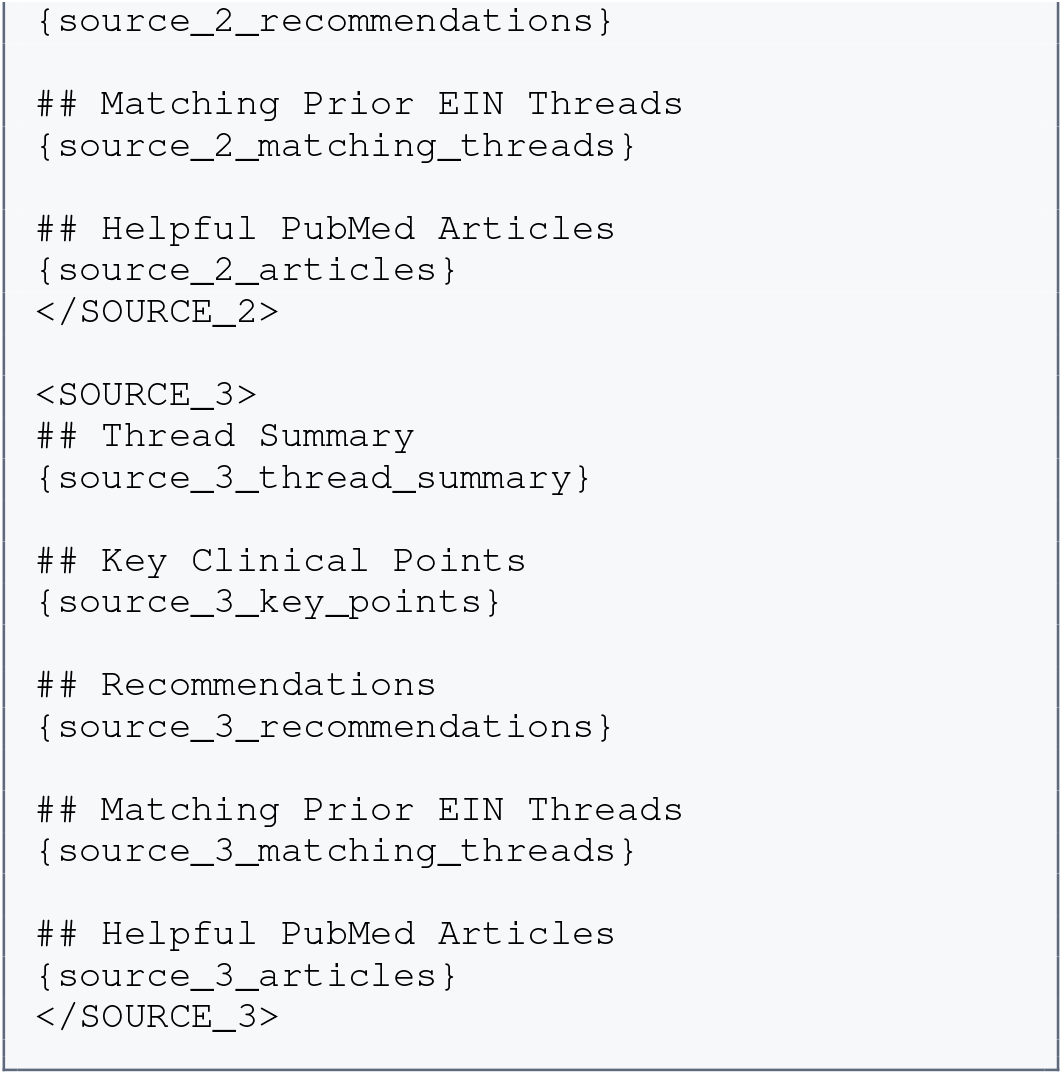
Ensemble Agent user prompt template.

Here, `{thread_id}` identifies the current EIN thread, and `{numbered_thread_posts}` contains the original post and replies in their conversation order. The remaining placeholders contain the corresponding sections produced by each of the three source agents. The Ensemble Agent returns Markdown rather than JSON because its output is the final clinician-facing summary.

## Outbreak Scenario-Based Analysis of LLM-as-Judge Scores

### Setup

We examined whether summary quality differed during major infectious-disease outbreak periods. Threads were assigned to an outbreak period using the date of their original post. We evaluated four periods for which sufficient data were available: the 2009 H1N1 influenza pandemic (April 2009– April 2010), the 2012 West Nile virus epidemic (March– December 2012), the 2012 pertussis epidemic (January– December 2012), and widespread hepatitis A outbreaks (August 2016–December 2019).

For each thread and system, we used the median score across repeated LLM-as-a-judge evaluations. We then computed the mean score within each time bin for four clinically salient dimensions: synthesis, recommendation actionability, recommendation appropriateness, and evidence calibration. The H1N1, West Nile, and pertussis plots use monthly bins; the longer hepatitis A period uses four-month bins to improve interpretability. Lines indicate mean judge scores and shaded regions indicate one standard deviation across threads in the corresponding time bin.

The figures include DeepSeek V1, OLMo V1, DeepSeek V2, and the Ensemble system. Formal paired Wilcoxon signed-rank tests compare DeepSeek V2 with each V1 baseline on the same threads. The Ensemble is shown descriptively because it is a downstream system that incorporates artifacts from the other candidate systems and uses a larger model; thus, its results should not be interpreted as an independent head-to-head model comparison.

## Results

Across outbreak scenarios, DeepSeek V2 generally achieved higher synthesis scores than both V1 baselines. This pattern was most consistent for synthesis and recommendation actionability, suggesting that the V2 retrieval-augmented workflow was particularly helpful for integrating multi-author clinical discussions into usable recommendations during outbreak-related conversations.

During the H1N1 pandemic (*n* = 125 paired threads), DeepSeek V2 improved synthesis relative to DeepSeek V1 (Δ = 0.30, *p* < 0.001) and OLMo V1 (Δ = 0.24, *p* = 0.005). It also improved recommendation actionability relative to DeepSeek V1 (Δ = 0.23, *p* = 0.004). Differences in recommendation appropriateness and evidence calibration were smaller and were not statistically distinguishable within this period.

During the West Nile epidemic (*n* = 87), DeepSeek V2 again improved synthesis relative to DeepSeek V1 (Δ = 0.23, *p* = 0.021) and OLMo V1 (Δ = 0.48, *p* < 0.001). It also improved recommendation actionability relative to OLMo V1 (Δ = 0.29, *p* = 0.002). During the pertussis epidemic (*n* = 113), the synthesis advantage remained evident relative to both DeepSeek V1 (Δ = 0.33, *p* < 0.001) and OLMo V1 (Δ = 0.52, *p* < 0.001), while recommendation actionability was higher than OLMo V1 by 0.34 points (*p* < 0.001).

The largest and broadest differences were observed during the hepatitis A outbreak period (*n* = 287). Compared with DeepSeek V1, DeepSeek V2 achieved higher synthesis (Δ = 0.39, *p* < 0.001), recommendation actionability (Δ = 0.26, *p* < 0.001), recommendation appropriateness (Δ = 0.24, *p* < 0.001), and evidence calibration (Δ = 0.17, *p* < 0.001). Compared with OLMo V1, DeepSeek V2 also showed higher synthesis (Δ = 0.49, *p* < 0.001) and recommendation actionability (Δ = 0.40, *p* < 0.001).

The Ensemble trajectory was generally at or near the highest mean judge scores across the four displayed criteria. This is consistent with the overall evaluation, in which the Ensemble performed especially well on synthesis, evidence calibration, and recommendation quality. However, these patterns are descriptive as the Ensemble receives outputs from the other systems, and its apparent advantage reflects the performance of the complete aggregation pipeline rather than an isolated effect of model choice.

Overall, the scenario-based analysis indicates that the V2 workflow preserved its advantages during multiple outbreak contexts, particularly for producing synthesized and actionable summaries.

